# A Poisson binomial based statistical testing framework for comprehensive comorbidity discovery across massive Electronic Health Record datasets

**DOI:** 10.1101/2021.07.14.21260532

**Authors:** Gordon Lemmon, Sergiusz Wesolowski, Alex Henrie, Martin Tristani-Firouzi, Mark Yandell

## Abstract

Discovery of comorbidities (the concomitant occurrence of distinct medical conditions in the same patient) is a prerequisite for creating forecasting tools for downstream outcomes research. Current comorbidity discovery applications are designed for small datasets and use stratification to control for confounding variables such as age, sex, or ancestry. Stratification lowers false positive rates, but reduces power, as the size of the study cohort is decreased. Here, we describe a Poisson Binomial based approach to comorbidity discovery (PBC) designed for big-data applications that circumvents the need for stratification. PBC adjusts for confounding demographic variables on a per-patient basis, and models temporal relationships. We benchmark PBC using two datasets, the publicly available MIMIC-IV; and the entire Electronic Health Record (EHR) corpus of the University of Utah Hospital System, encompassing over 1.6 million patients, to compute comorbidity statistics on 4,623,841 pairs of potentially comorbid medical terms. The results of this computation are provided as a searchable web resource. Compared to current methods, the PBC approach reduces false positive associations, while retaining statistical power to discover true comorbidities.

## Introduction

Comorbidity refers to the concomitant occurrence of distinct medical conditions in the same patient^1^. Comorbidities can occur together, or sequentially across the patient’s medical history. Exploring these temporal connections offers additional insight into disease progression and disease associations, and promises improved predictive tools for evidence-based medicine^2–5^. Traditionally, comorbidities have been discovered manually, through human chart review, literature search, and clinical knowledge^6^. For example, the authors of the Charlson comorbidity index^7^ selected comorbid diagnoses based on manual chart review for a 559-patient cohort. Likewise, the well-known Elixhauser Comorbidity index, was compiled through review of published studies identifying comorbid conditions^8^.

Large collections of Electronic Health Records (EHRs) present promising new opportunities for comorbidity discovery. However, manual review of millions of EHR records in search of comorbidities is infeasible, and *ab initio* means for discovery and temporal ordering of comorbidities using large collections of EHRs is an area ripe for innovation. Current computational approaches to *ab initio* comorbidity discovery use statistics such as risk ratio, odds ratio, comorbidity-score^9^, propensity score or ϕ-correlation to measure effect size. P-values are obtained using Fisher’s exact test (or hypergeometric), χ² test, or binomial test^10–14^. All of these approaches rely on an assumption that each member of the population has a disease probability equal to the population incidence rate. Confounding variables such as age and sex are controlled for by sub-setting the data, a process termed s*tratification*, or alternatively through use of matched case-control cohorts^14, 15^. Both of these approaches control for confounders, but at the expense of statistical power, because they necessarily reduce sample size. One approach to overcoming this intrinsic limitation is to aggregate massive collections of EHRs, but as we show, even millions of records are too few to explore comorbidities associated with rare diseases when controlling for multiple confounding variables.

PBC models the effects of confounding variables, allowing every sample to be personalized, resulting in improved statistical power compared to stratification. Briefly, PBC uses logistic regression to model how each patient’s demographics impact his or her probability of having a medical term. These personalized probabilities are then used to calculate pairwise expectations and p-values under the Poisson binomial distribution, rather than the hypergeometric and binomial distributions that are used for Fisher’s exact test, and the *χ*² test, respectively. Moreover, with minor modification, this approach can also be used to temporally order comorbidities and to determine the significance of directionalities. As we demonstrate, use of the Poisson binomial is a significant advantage, because it obviates the need for stratification. The result is increased power for discovery, which we leverage to explore the relationships among diagnoses, medical procedures and medications.

Alongside the need for improved statistical methods, tools are also needed to browse, search, and visualize the network of comorbid medical terms discovered in big-data applications. In a manner similar to *Siggaard et al.*^14^, we provide a browser-based query engine for navigating these comorbidities and their temporal relationships within the University of Utah Hospitals system [https://pbc.genetics.utah.edu/lemmon2021/pbc-utah].

In what follows, we describe PBC, explore its behavior, and benchmark its performance using the contents of the University of Utah Health system, and the publicly available MIMIC-IV dataset^16^. For brevity’s sake, we will refer to co-occurring medical diagnoses, procedures and medications using the single blanket term, comorbidity. We demonstrate how PBC can be used to transform massive EHR datasets into a temporal dependency graph for large-scale *ab initio* discovery of comorbid relationships and investigations of disease progressions.

## Results

### Modeling the effects of confounders using logistic regression

We collected records for 1.6 million patients, encompassing 50 million visits and 150 million diagnosis (DX), procedure (PX) and medication (RX) codes from the University of Utah Electronic Data Warehouse (EDW). For the proof of principle analyses presented here, diagnoses were converted from ICD9^17^ and ICD10^18^ diagnosis codes to Clinical Classification System^19^ (CCS) multi-level diagnosis codes. CPT^20^ provider billing codes were converted to CCS multi-level procedure codes. We include both leaf nodes and internal nodes so that the researcher can discover more specific comorbidities (e.g “CCS 2.1.1: cancer of colon”) as well as more general comorbidities (e.g. “CCS 2.1: colorectal cancer” or “CCS 2: neoplasms”). Medications were coded using RxNorm concept unique identifiers (CUIs)^21^. This procedure reduced the number of distinct medical terms to 1007 DX, 259 PX and 1775 RX codes. We also collected demographic information, including sex, race, ethnicity, insurance class, age and length of medical records.

A logistic regression model (LRM) was determined for each DX, PX and RX term. The LRM includes demographic information for each patient (age, gender, ancestry, ethnicity, insurance type) and EHR exposure (the length and density of a person’s medical record). Since medical terms are included or excluded from year to year, and coding practices vary over time, we also include the date of the patient’s last visit as a control for this effect. The complete list of features is described in **Table S1** and **Figure S1**. The response variable is whether each patient has the term in their medical record. Note that we do not model recurrence - in this analysis we consider only the first instance of a term in a patient’s medical history. We use these LRMs to estimate for each medical term, each patient’s *a priori* personalized probability of having that term in their medical record.

**Table 1.**
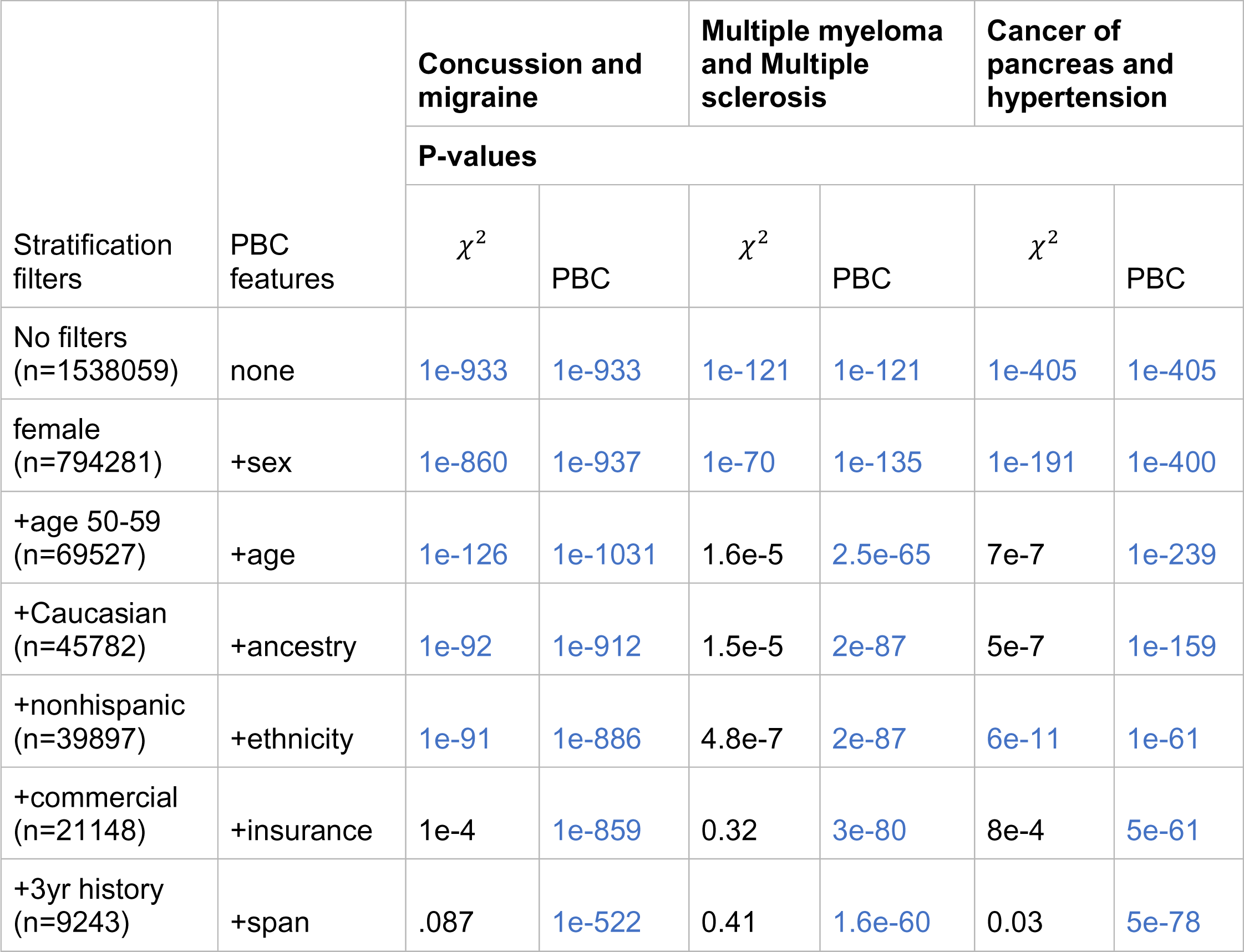
PBC retains power as features are added; stratification loses power. Shown are 3 established comorbidities from the medical literature: concussion and migraine (41), multiple myeloma and multiple sclerosis (42) and cancer of pancreas and hypertension (43). Comorbidities passing a Bonferroni corrected alpha threshold of **1.08e-8** are colored blue. As features are added to the stratification criteria, sample size shrinks and statistical significance is lost. Rather than controlling for confounding variables, PBC models their effects. Thus, for PBC sample size remains constant at 1,538,059 and statistical significance is preserved.

We used a regularized regression model under the assumption of collinearity in the confounding features. However, there is no requirement for such an assumption; for example, Neural networks could be used in place of LRM, so as to better capture nonlinear relationships between variables. L1 and L2 penalized logistic regression include a value *C* that prevents overfitting by penalizing large coefficients. Smaller *C*-values specify stronger regularization (e.g., stronger prevention of over-fitting). To determine the optimal C-value for each LRM, it was necessary to choose a score function for LRM evaluation. We experimented with a number of standard and custom score functions as described in supplemental “Math.pdf”. We optimize C for each LRM using stratified 3-fold cross validation. Grid search is used to evaluate C-values in the set {*{10^-14^, 10^-13^, 10^-12^, …, 10^12^, 10^13^, 10^14^}*. **Figure S2** top panel shows boxplots of the scores reported by each of these score functions. We use entropy as a measure of the ability of a score function to differentiate model quality. From those score functions achieving high entropy, we evaluate the distribution of C-values (**Figure S2** bottom panel). We choose J_*cutoff*_ for all downstream analysis because it includes fewer outliers than the other methods examined. J_*cutoff*_ is based on Youden’s J statistic^22^, only rather than a 50% probability threshold, the classification threshold is determined empirically so that the total number of predicted positives is equal to the actual count of positives.

We train LRMs using stratified 3-fold cross validation, evaluated using J_*cutoff*_. Under L1 penalized logistic regression, model features can be unselected by setting their coefficients to zero. **Figure 1** summarizes how often each demographic feature is included in the trained LRMs. Age at last visit, number of visits, number of terms, and length of medical record were grouped together as “EHR exposure”. Patients may be seen at a non-university clinic, may move in or out of the state over the years, or may have differing proclivities toward visiting the doctor. EHR exposure is an attempt to control for these effects. As **Figure 1** makes clear, EHR exposure is always important in predicting whether a patient has a particular medical term.

**Figure 1.**
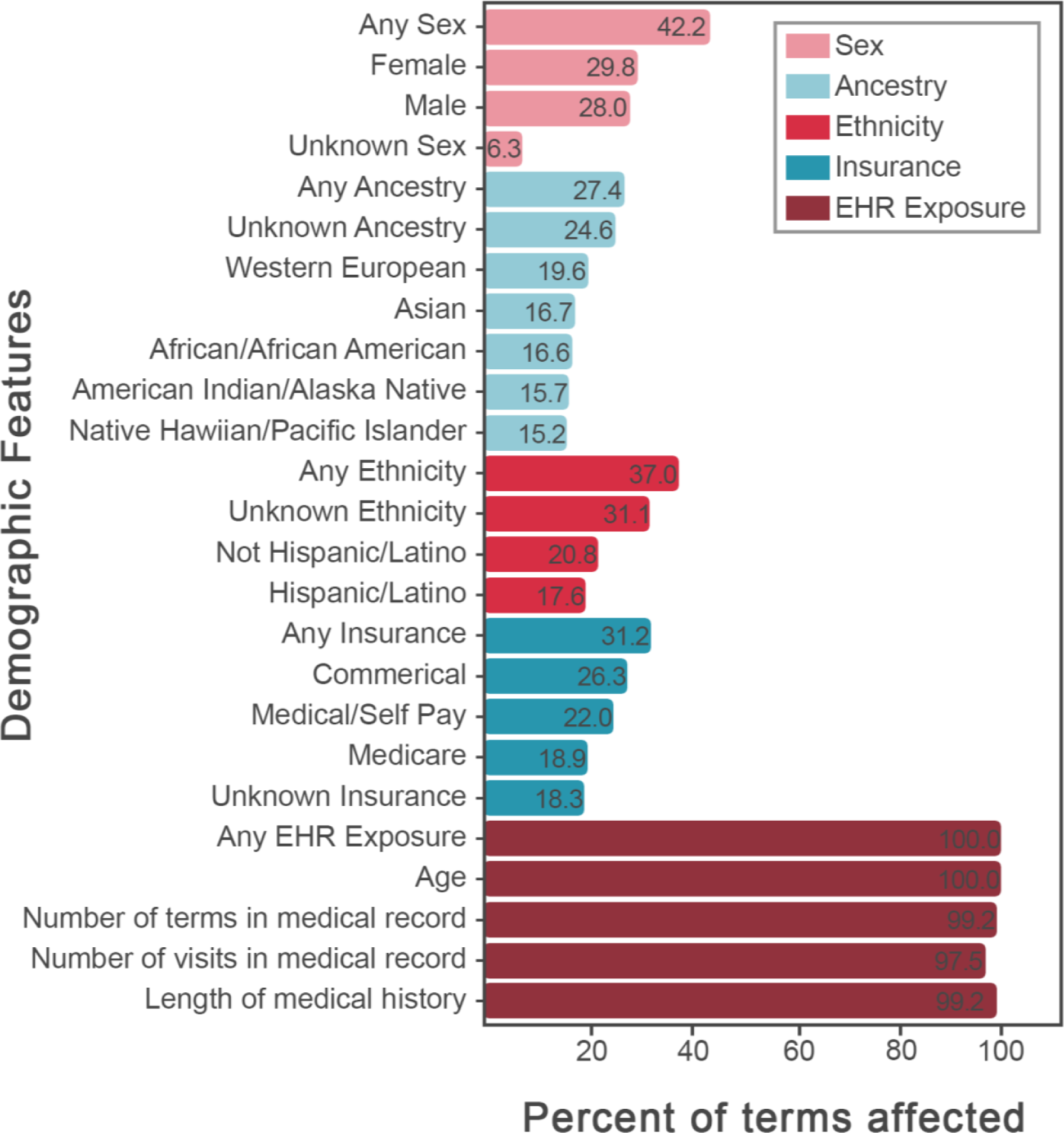
Feature selection by L1 regularization. Percent of medical term logistic regression models that include each demographic feature. For example, EHR exposure, i.e., the length and density of a person’s medical history, is an important predictor for every medical diagnosis, procedure and medication.

### PBC retains power as features are added and as sample sizes are reduced

The binomial distribution models the discrete probability of the number of successes in N independent experiments each with probability P. A naïve approach to comorbidity analysis assumes the probability of seeing term 1 and term 2 (*P*_𝓉1,𝓉2_) in a medical record is the product of the population incidence rates for terms 1 and 2. Knowing the number of patients with both terms 1 and 2 in their medical record, one can calculate a comorbidity p-value using the binomial test of statistical significance. However, because this method does not adjust for demographic factors, the p-values they generate can be driven by effects such as age and sex. Thus, a common approach in comorbidity literature is to stratify the population by age and sex, and then calculate the binomial p-value for each stratum^7^. In contrast, the PBC approach uses the Poisson binomial distribution to calculate p-values for each term pair. The Poisson binomial distribution is a generalization of the binomial distribution in which every trial/sample (i.e., patient) has a different probability of “success” (i.e., having both terms in their medical record). The probability of an individual patient having both terms in their medical record is calculated as the product of per-patient per-term probabilities generated from corresponding LRMs described above.

**Figure 2** explores the relationships between comorbidities discovered by PBC versus stratification as a function of increasing numbers of demographic features. As can be seen, the stratification approach rapidly loses power as more criteria are added to the stratum filter. In contrast, by modeling demographic features, PBC maintains power.

**Figure 2.**
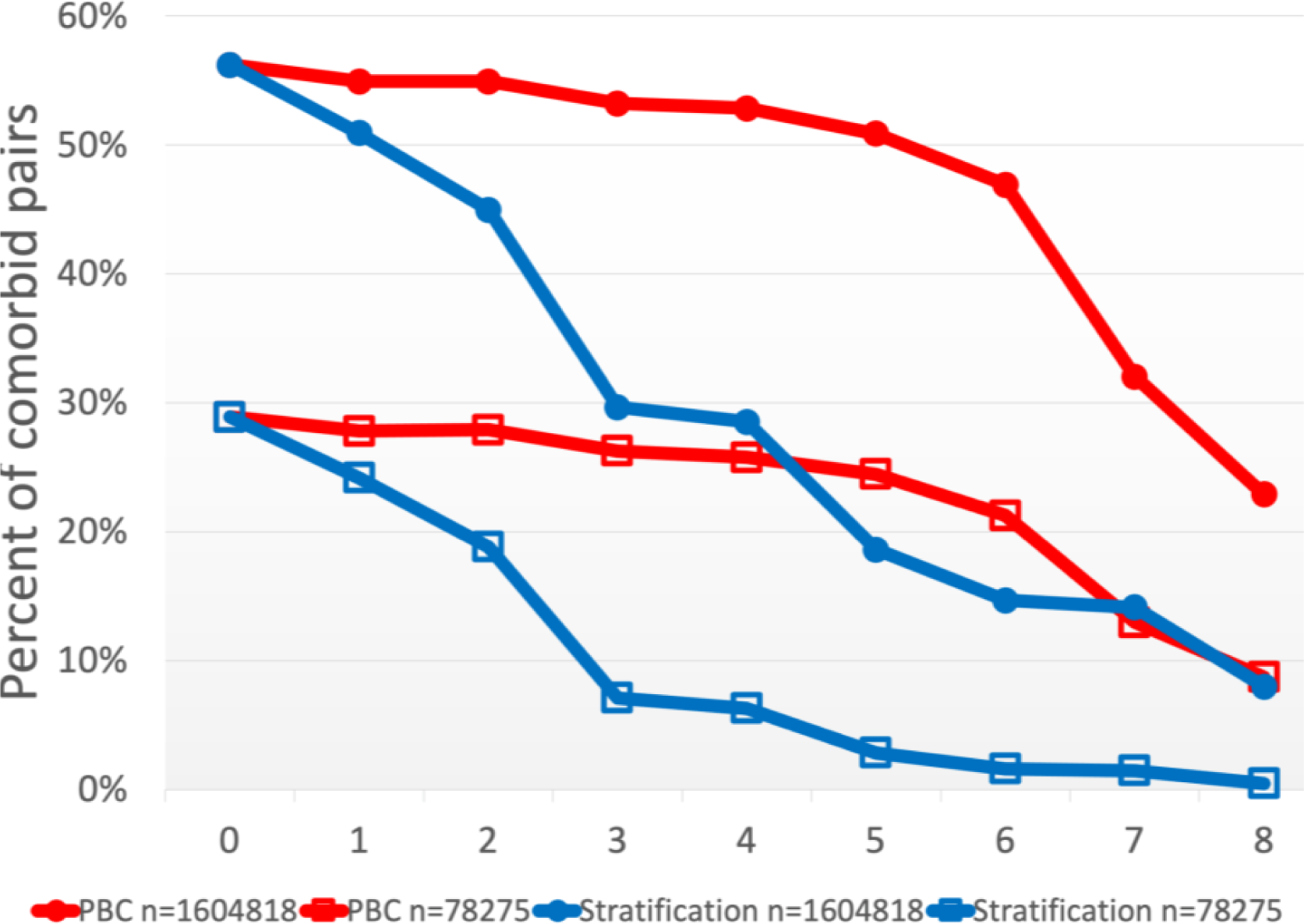
PBC maintains power for discovery by modeling the effects of confounding variables. We calculate the percent of significantly co-occurring pairs of medical terms (p<1.08e-8) using either PBC (blue lines) or stratification (red lines) for two different sample sizes (Entire EHR corpus [1.6 million individuals, filled circles], and a 78,275 patient sample [open squares]. Moving from left to right we introduce additional features to the PBC approach and additional filters to the stratification approach. The X-axis numbering corresponds to the following features/filters: 0: no features; 1: race / African American; 2: sex / Female; 3: Age / 50-59; 4: Ethnicity / nonhispanic; 5: Insurance / Commercial; 6: Span / at least 2 years; 7: Number of visits / at least 3 visits; 8: Date of last visit / at least as recent as Jan 2018. The figure highlights 2 important trends. First, the number of significant associations decreases as a function of smaller datasets. Second, controlling for specific features markedly reduces the number of recovered comorbidities for the stratification approach (an effect further exacerbated by reducing the initial cohort size), while preserving significant comorbidities using PBC.

**Table 1** compares the power of stratification and PBC to detect three well known comorbidities using our EHR corpus. As in **Figure 2**, we compare the p-values generated by sequentially adding confounding variables. Notice how stratification dramatically lowers the strength of p-values as a function of the size of the stratum. This same behavior is illustrated globally in **Figure 2**. Even with millions of EHRs, controlling for more than a few confounding demographic variables leads to strata that are too small to achieve statistical significance. By modeling the effects of multiple confounding variables, PBC retains statistical power to identify comorbidities.

**Table S3** presents five well known comorbidities of breast cancer^23–28^. Stratification by age and gender deflates the strength of all p-values, and as a result they fall below the Bonferroni corrected significance threshold (**Table S3** column “binomial female 50-59”). In contrast, by explicitly modeling age, gender, race, ethnicity, insurance type, and EHR exposure, PBC retains power to capture these true positive associations.

**Table 2.** PBC identifies comorbidities specific to underrepresented minorities even when data is limited. Here we compare the stratification-based approach with PBC as regards ability to identify a known comorbidity: sickle cell anemia (SCA) paired with malaise and fatigue. P-values passing a Benjamini-Hockberg corrected alpha threshold of **1.0e-6** are colored blue. Without modeling the effects of confounding variables, both approaches identify the association. But because ancestry is a key determinant of risk for SCA, we need to control for this confounding variable to determine whether malaise and fatigue is an actual symptom of sickle cell anemia or whether the connection is being driven by ancestry. Filters applied under stratification lead to samples too small to detect this association. In contrast, not only does PBC detect the comorbidity, but the strength of the association increases as confounders such as ancestry, ethnicity and age are included in the model.

| Stratum filters | PBC features | Sickle Cell Anemia paired with Malaise and Fatigue |  |  |
| --- | --- | --- | --- | --- |
| | | Stratum Pair count | $\chi^2$ p-value | PBC p-value |
| No filters (n=477,070) | no features | 19 | 4e-8 | 4e-8 |
| Caucasian (n=276,496) | ancestry | 5 | 0.002 | 2e-8 |
| African American (n=7,035) | ancestry | 9 | 2e-5 | 2e-8 |
| +Nonhispanic (n=6,039) | +ethnicity | 8 | 2e-4 | 1e-8 |
| +Female (n=2,789) | +gender | 7 | 1e-4 | 9e-8 |
| +50-59 (n=302) | +age | 3 | 0.004 | 3e-19 |

**Table 3.** PBC is the only comorbidity search tool that takes a high-resolution approach. CytoCom and comoR are no longer available; comoRbidity and Comorbidity4j failed to scale to datasets of our size. In addition to scaling to handle arbitrarily large EHR datasets, PBC models the effects of demographic information rather than relying on stratification.

| Package | CytoCom | comoR | comoRbidity | Comorbidity4j | DTB* | PBC-web |
| --- | --- | --- | --- | --- | --- | --- |
| Release date | 2014 | 2014 | 2018 | 2019 | 2020 | 2021 |
| Relative Risk<sup>†</sup> | ✓ | ✓ | ✓ | ✓ | ✓ | ✓ |
| ϕ-correlation<sup>†</sup> | ✓ | ✓ | ✓ | ✓ | | ✓ |
| Comorbidity score<sup>†</sup> |  |  | ✓ | ✓ |  | ✓ |
| Odds ratio<sup>†</sup> |  |  | ✓ | ✓ |  | ✓ |
| Fisher's Exact test<sup>†</sup> |  | ✓ | ✓ | ✓ |  | ✓ |
| $\chi^2$ or Binomial test | | | | | ✓ | ✓ |
| Poisson Binomial<sup>‡</sup> |  |  |  |  |  | ✓ |
| Inverse comorbidities |  |  |  |  |  | ✓ |
| Temporal directionality |  |  | ✓ | ✓ | ✓ | ✓ |
| Interactive network of comorbidities | ✓ |  |  |  | ✓ | ✓ |
| Arbitrarily large datasets |  |  |  |  | See note<sup>§</sup> | ✓ |
| Dealing with confounders | stratify | no support | stratify | stratify | matching | Logistic regression |
| Platform | Cytoscape | R package | R package | Java/website | Publicly available website | Publicly available website |

\*Disease Trajectory Browser
<sup>†</sup>Statistics that rely on population incidence rate
<sup>‡</sup>Statistics that rely on per-person per-term probabilities
<sup>§</sup>Authors apply a pre-filtering step because calculating all pairs of comorbidities is too computationally demanding

The complete set of comorbid term pairs discovered by PBC can be visualized using network analysis. In supplemental **Figure S3**, we use the minimum description length algorithm^29^ to perform clustering of pairwise comorbidities by p-value strength. Terms with similar patterns of comorbidities are closer together in the network. To illustrate this, we annotated selected comorbidities discovered within each cluster. A literature search confirmed that these labeled comorbidities represent existing clinical knowledge (see corresponding citations in **Table S7**). **Figure S3** provided the motivation to produce an interactive tool for querying, exploring and extracting information from the comorbidity network. In order to provide better means to navigate this complex network we developed a browser-based tool for exploration of comorbidities, discussed below.

### PBC identifies comorbidities unique to underrepresented minority groups

PBC can also capture true comorbidities hidden within mixed populations. For instance, consider Sickle Cell Anemia, a disease that affects 1 in 365 African American newborns and 1 in 100,000 newborn Caucasians in the United States^30^. Malaise and fatigue, while common to many disorders, are among the most common symptoms of Sickle Cell Anemia (SCA)^31–33^, and usually manifest in an age dependent manner. **Table 2** compares five comorbidity p-values. Row 1 presents p-values calculated using all data. While the true comorbidity is discovered, we cannot say with certainty if the relationship is a true comorbidity or simply driven by a third confounding variable (e.g. ancestry). The following rows present X2 p-values after stratifying by ancestry, ethnicity, gender, and age and PBC p-values after including these same features in the regression model. Stratification fails to find a significant comorbidity between SCA and maliase/fatigue once the data is partitioned by ancestry. The rarity of Sickle Cell disease in Caucasians and the small sample size for African Americans within Utah’s EHR corpus make detection of this comorbidity difficult. PBC, in contrast, discovers the comorbidity. In fact, the additions of ancestry, ethnicity, and age each increase the strength of the association between SCA and malaise/fatigue.

### Application of PBC to a publicly available dataset

Because the University of Utah data used in this study cannot be shared publicly, as it includes protected health information (PHI), we also demonstrated the general applicability of PBC by applying it on the publicly available MIMIC-IV dataset^16^. This dataset includes 248,714 patients with associated ICD10 diagnosis or procedure codes. A total of 5,363,338 ICD9 and ICD10 diagnosis and procedure codes were converted to multi-level CCS codes. These include 725 distinct CCS diagnosis codes and 395 distinct CCS procedure codes. We repeated our experiments on this dataset, training a logistic regression model for each diagnosis and procedure code, and calculating comorbidities using the Poisson binomial distribution. Although the MIMIC-IV data is missing much of the detail available in the University of Utah dataset, the PBC results generally mirror those of the Utah dataset (**Figure S4**). Regression features are shown in **Figure S4**, top panel. In addition, because a random offset is added to each patient’s admission dates, it is not possible to control for the changes in use of various billing codes over time. Despite these limitations, the deployment of PBC on the MIMIC-IV public dataset further illustrates how PBC retains statistical power to identify comorbid relationships that are lost by stratification. Tables 1 and 2 are replicated on MIMIC-IV data as supplemental tables S4 and S5. Co-occurrence and directional comorbidities discovered within MIMIC-IV data can be queried at the following link: https://pbc.genetics.utah.edu/lemmon2021/pbc-mimic.

### Temporalized P-values allow for understanding of disease progression

The PBC approach can be extended to provide temporalized (or directional) p-values across pre-specified time windows (see Methods for details). The inclusion of a direction window is necessary for several reasons. On short time scales, the order of appearance of diagnostic codes is an unreliable indicator of which condition actually preceded the other in the patient. The development of underlying disease, the relevant signs and symptom, the provider arriving at a given diagnosis, and the eventual recording of the said diagnosis might follow staggered paths that have little or no relevance when viewing the data in a time-slice of less than 30 days or so, thus for many analyses it is probably best to treat these events as contemporaneous. However, in some cases a short window size is optimal for capturing a comorbidity.

Consider the following example. PBC reports the following p-values for the diagnosis/procedure pair amputation of lower extremity → postoperative infection: within 30 days = 1e-3663, greater than 30 days = 1e-24, greater than 90 days = 1e-6, greater than 365 days = 0.86. It is clear that shorter window sizes better capture the increased risk of infection after amputation, as one would logically expect. Thus, the window size must be informed by clinical knowledge and research objectives. In this manuscript, we examined associations based on a 90-day window. Using our website (see below) a user can also query additional window sizes (30 days, 365 days and 730 days).

**Table S6** presents specific directional associations discovered in an *ab initio* fashion by PBC and supported by clinical knowledge. For instance, Milrinone is prescribed to patients awaiting heart transplant^34^. The tendency of type 2 diabetics to develop chronic kidney disease is well known^35, 36^. HIV induced immunocompromisation often leads to pneumocystis^37^ and obesity is a known risk factor for hypertension^38^.

### A web-based resource for comorbidity research

Among 4,623,841 pairs of medical terms in our collection of 1.6 million EHRs, we identified 3,311,830 comorbidities co-occurring within a 90-day window, and 1,969,941 temporally directed ones, acting over a time period greater than 90 days. All associations meet a Bonferroni significance threshold of 1.08e-08. The result is a highly-connected network of comorbid diagnoses and associated procedures and medications based on the University of Utah EHR database.

In response to the size and complexity of these ‘big-data’, we have created browser-based means to navigate, query and explore them (https://pbc.genetics.utah.edu/lemmon2021/pbc-utah). A screenshot highlighting the functionality of the browser is shown in **Figure 3**. The site allows the user to search for relationships between any pairwise diagnosis, procedure or medication. The result is a searchable table of all other DX, PX, and RX codes along with statistics about the connection to the query term. These statistics include the counts, expectation and p-value of association after adjusting for confounders shown in **Figure 1**. We use two-sided p-values, so that “less than expected” associations are also discoverable. For comparison, we provide both χ^2^ p-values and G-test p-values.

**Figure 3.**
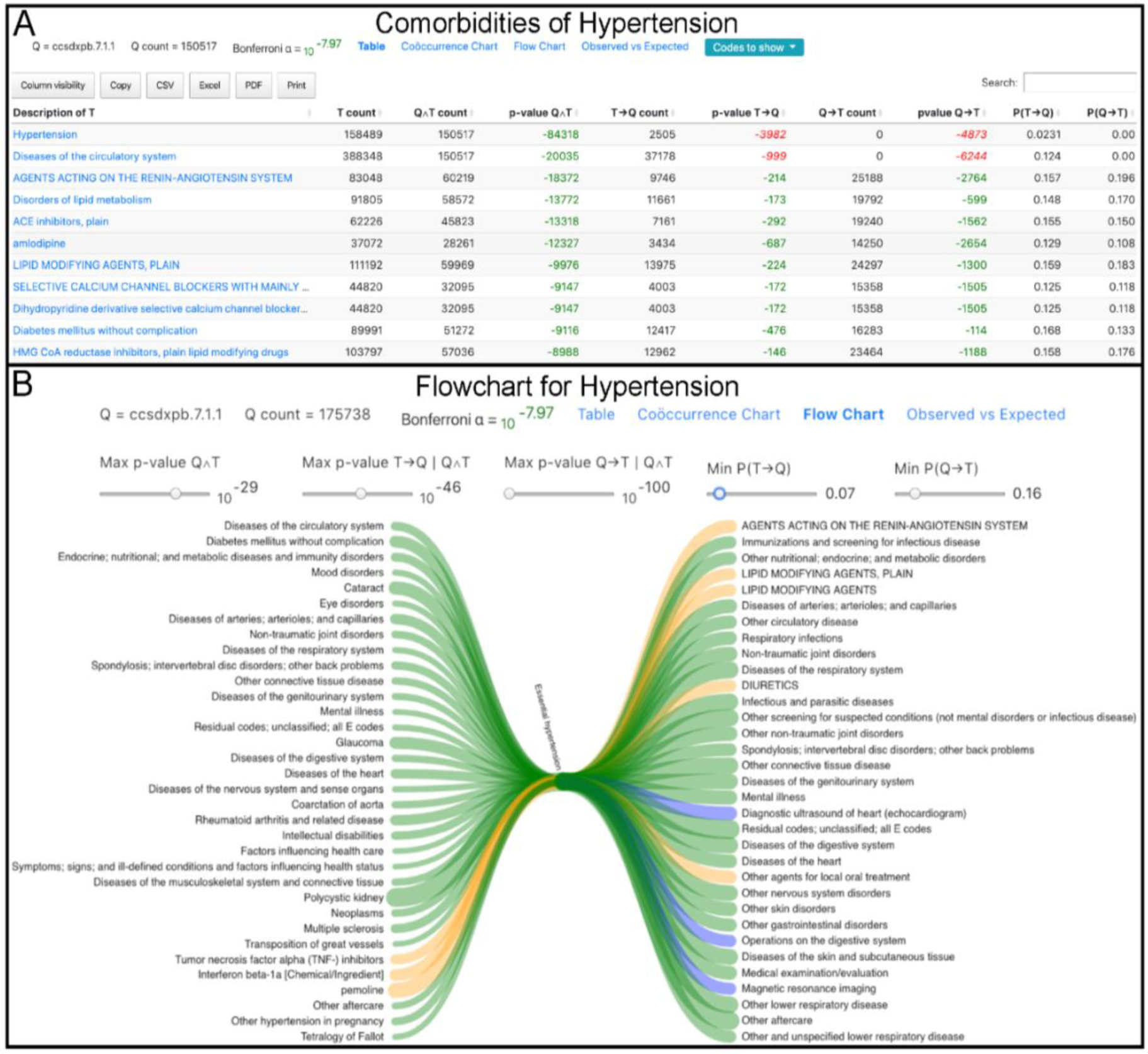
Screenshots from PBC web. “Q” refers to the query term, in this case hypertension. “T” refers to the term possibly comorbid with Q. **Panel A**, Code prefixes in the first column can be deciphered as follows: ccs = “clinical classification system”, dx = diagnosis, px = procedure, pb = provider billing (we omit hospital billing codes in this figure), cui = RxNorm concept unique identifier. P-values that pass the Bonferroni corrected significance threshold are colored green or red. Green indicates the relationship occurs more often than expected. Red indicates less often than expected. The last two columns represent flow rates which indicate the actual percent of patients in our database that transit from one term to the next over time. **Panel B**, Terms that significantly precede or follow hypertension (separated by at least 90 days) are shown to the left and right of hypertension respectively. Green connections are diagnoses, blue connections are procedures and orange connections are medications. The thickness of the connection relates to the flow rate – the percent of patients that flow through the given path.

In addition to patient lifetime co-occurrence p-values, we provide within window, and out of window directional p-values with a selectable window size (30, 90, 365, or 730 day). We also provide statistics on effect size, including relative risk, odds ratio, and “flow rate” which is the percent of patients coded with term 1 who later are coded with term 2. The “Flowchart” view puts the query term in focus and shows the terms that tend to precede and follow the query term to the left and right respectively. The user can filter by p-value strength and by flow rate.

A slice of that network is shown in **Figure 3**. This figure contains a “flow chart” view for essential hypertension. By switching out the central node, an investigator can step through the temporalized network of diagnoses, procedures and medications. The investigator can further filter by effect size to find co-occurrences that are both significant and prevalent.

### Comparison with other published comorbidity tools

**Table 3** compares functionalities provided on our comorbidity website with those found in other published comorbidity discovery tools. To the best of our knowledge, our website is the only available public resource of its kind that considers the individual risk profile for each patient having each medical term. Additionally, our approach (and website) captures inverse comorbidities (terms occurring together less often than expected) and models temporal relations between pairs of terms.

The R package “comoRbidity” was published April 2018^10^. We installed the package, and reformatted our data to fit the required specifications. Given our input of 150,598,377 CCS and RxNorm codes, the “comoRbidity” package consumes all available RAM (we used a Linux server with 504 GB of RAM) and fails to complete. We tried to acquire CytoCom^11^ and comoR^12^, however the corresponding author has indicated that these projects are no longer maintained nor available for download. The Java package “Comorbidity4j” was published January 2019^13^. We installed Comorbidity4j and attempted to compute on our full dataset. The application warned against calculating comorbidities for more than 300,000 pairs of terms (774 distinct terms). After several hours the calculation times out - having allocated 50.2 GB of RAM. In contrast our method uses a maximum of 174 MB of RAM and has no upper limit on the number of patients, visits, or unique terms.

The Disease Trajectory Browser (DTB) allows navigation of temporal relationships among medical records of 7.2 million Danish patients^14^. Direct comparison between our results is not possible since our groups have access to different EHR datasets, however we can consider differences in methodology. DTB measures effect size using relative risk, significance is measured using the binomial test, and confounders are controlled for using case/control matching. For comparison, on PBC-Web, we provide relative risk estimates and χ^2^ p-values (which are a close approximation to the binomial test p-values). As described above, our approach models patient features rather than controlling for them through stratification or case/control matching.

## Discussion

Although the term comorbidity is often used to denote significant associations between medical outcomes, (e.g., hypertension and heart attack), the concept is easily extended to include associated variables, such as medical procedures and medications. For brevity’s sake, in what follows, we refer to statistically significant associations among these collective variables as comorbidities.

Comorbidity discovery is a feature discovery/selection process, and it is important to distinguish it from outcomes prediction. Before one can understand and predict a medical outcome, one must first decide which prior diagnoses, medical procedures and medications are germane to the outcome of interest. Discovery of comorbidities is thus a prerequisite for downstream outcomes research and for creating forecasting tools^39–42^, but is logically separate from them.

Big data offer many opportunities and challenges for comorbidity discovery. One limitation imposed by data size is that morbidity discovery is necessarily pairwise; hence the term comorbidity, as opposed to multimorbidity discovery. Tools for multimorbidity-based discovery^43^ are necessarily limited in scale due to computational constraints, considering for instance, 34 disease clusters^44^. In contrast, we have calculated pairwise comorbidities among 37,997 ICD10 diagnosis codes.

Commonly used statistical approaches to *ab initio* comorbidity discovery are hindered by the assumption that every member of the population (or stratum) has a disease probability equal to the population (or stratum) incidence rate. As we have explained, stratification is commonly used to subset EHR collections to meet this requirement; but stratification necessarily reduces sample size and statistical power (c.f. **Figure 2**). In practice, stratified data quickly become limiting, even for very large datasets, as inclusion criteria grow more complex. This problem is exacerbated for *ab initio* approaches aimed at simultaneous discovery of comorbid relationships among thousands of diagnoses, procedures and medications. This process necessitates many millions of statistical tests; the requirement for multiple testing corrections mean that statistical power is of paramount importance.

Our motivation in developing PBC was to overcome the need for stratification, while still achieving high accuracy and statistical power, so as to allow discovery of comorbidities of rare diseases, using small datasets, and *ab initio* discovery using very large EHR collections. Our results document the efficacy of PBC for achieving these ends. Still, it is important to bear in mind, that the comorbidities it discovers do not necessarily indicate mechanistic relationships. For instance, two diagnoses may both be driven by smoking, but since smoking was not included logistic regression model, we cannot say anything about smoking as a potential driver (cause) of the relationship. Thus PBC, like all existing methods in this domain, cannot with certainty assign comorbidities to one of the four etiological models described by Valderas et al^1^; it can only say that the relationship is not due to factors included in the logistic regression model.

Our hope is that PBC will provide an effective solution for a foundational step for outcomes research. The curse of dimensionality is a well-known phenomenon in which training a predictor with too many features can lead to higher error rates^45, 46^. Considering there are about 69,000 ICD10 diagnosis codes, 70,000 ICD10 procedure codes, and 350,000 RxNorm CUI codes, dimensionality reduction is necessary for effective machine learning on EHRs. By discovering which variables influence which outcome, PBC can reduce dimensionality and facilitate the creation of downstream tools for outcome predictions. Thus, PBC’s role in feature selection becomes clear. For a given clinical outcome, PBC can produce a manageable set of pairwise associations which become the inputs for training predictive models of disease.

It is important to note the limitations inherent in the use of data from a single EHR for comorbidity discovery. Data from a single EHR represents a non-random sampling of the general population and PBC does not model this sampling bias. Billing practices can vary within hospital systems - for instance, between inpatient and outpatient services and between provider billing and hospital billing. Hospital billing is performed by medical billing specialists, whereas provider billing is performed by clinicians. In this paper, we have restricted our analysis to provider billing terms. Clinical notes provide a still richer, more nuanced source of data and may more accurately describe a patient’s medical condition. Clearly application of PBC to the outputs of Natural Language Processing (NLP) tools will be a fruitful path for future research.

## Conclusion

Capobianco and Lio^47^ present a vision for comorbidity discovery and analysis that is multi-disciplinary and enabled by dynamic networks, with time as a key component in explaining disease relationships. We share this vision, and our PBC method directly addresses the challenges for creating a scalable network-based approach that can (1) dynamically adjust for confounding demographic variables and (2) model temporal relationships in large, complex EHR datasets. However, comorbidities do not exist as isolated pairs, rather they combine in a conditionally dependent manner to create a complex web of influence on any given outcome. While PBC is powered to discover that web by identifying the major drivers of a particular outcome, determining the joint contributions of conditionally dependent variables on that outcome requires a separate computational machinery. Bayesian networks^48–49^, for example can be used to compute the joint contributions of multiple conditionally dependent variables (so-called multimorbid calculations), providing fully explainable patient outcome predictions.

Obtaining a global overview of comorbidity and disease progressions across a major research hospital network is as difficult as it is desirable. We offer the PBC web-browser as a first-generation navigation tool for this new domain of EHR database visualization. Our hope is that the PBC website will provide a community resource for outcomes research, laying the foundation for improving current comorbidity-based outcomes tools, creation of new ones, and, more generally, fueling healthcare discovery for improved care.

## Methods

### University of Utah medical records

The University of Utah maintains an Electronic Data Warehouse (EDW) – a central storage and search facility for all data collected from all university hospitals and clinics, and all departments and specialties. SQL queries were composed to the following information: (1) medical record number, sex, race, ethnicity, and age for each patient; (2) list of patient visits, along with visit date, and medical terms associated with each visit, including diagnostic codes, procedure codes, and medications ordered. Data were deidentified.

We collect ICD9^17^ and ICD10^18^ diagnosis codes CPT procedural codes^20^ and RXNorm^21^ medication codes (“concept unique identifiers”) from University of Utah electronic medical records. ICD and CPT diagnosis and procedure codes were mapped to the hierarchical Clinical Classification System (CCS)^19^. CCS codes allow for more powerful statistics at the expense of concept resolution. After mapping to CCS, we retain 1007 distinct diagnosis codes and 259 distinct procedure codes. These codes include both internal nodes and leaf nodes in the hierarchical CCS tree. In all, we collected records for 1.6 million patients, 50 million visits and 150 million diagnosis (DX), procedure (PX) and medication (RX) codes.

Counts of EDW patient demographics are displayed in **Table S1**. **Figure S1** displays how our data is distributed by gender and age decade. **Figure S1**, panel B shows how the length of patient medical records (in years) is distributed. Note that these lengths are limited by the history of electronic data collection at the University of Utah that began in the early 2000s but started to ramp up around 2009 and has since increased rapidly. Thus, we see the 95^th^ percentile for medical history length is around 12-15 years for most age bins. **Figure S1** panels C and D show how the number of visits and the number of terms in a medical record trend with age. In almost all decades, women have more medical visits and medical terms than men, though this effect is most pronounced between 20 and 50.

### Logistic regression for person-term probabilities

The initial step in comorbidity analysis is to ascertain the probability of a given term being found in a given person’s medical record. A naïve approach could assign everyone the same probability based on the term’s frequency in the database or within each age-gender strata as seen in other methods.

Our approach involves developing a logistic regression model for each term. The independent features, *X*, are the list of persons in the EHR along with their gender, race, ethnicity, financial class and risk exposure. Risk exposure includes the age of a person at the time of their last visit as well as the length and density of their medical record. Length is defined as the number of days between first visit and last visit, while density is approximated by the number of visits within a medical record. The dependent outcome, *y*, is a binary vector indicating whether each person has the term in their medical record. The value *C* is the inverse of regularization strength in L2 penalized logistic regression. Smaller values of *C* indicate stronger regularization. The coefficients are determined by minimizing the following loss function, where *β* represents the coefficients and *c* is a constant (see Scikit-learn documentation^50^ for a more complete discussion of logistic regression):

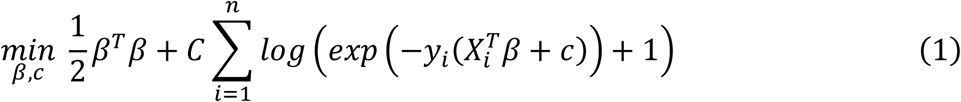

For each term, stratified 3-fold cross-validation is used to determine the optimal value for *C within the set {10^-14^, 10^-13^, 10^-12^, …, 10^12^, 10^13^, 10^14^}*. Cross-validation relies on a scoring function to assess the accuracy of logistic regression, given differing values for *C*. We evaluated standard and custom score functions based on their ability to differentiate logistic regression results with differing *C* values (see Figure S2).

The above approach resulted in logistic regression models for each term, capable of predicting the probability that a given person has a given term. For rare terms, we find that the probabilities output by logistic regression may not sum up to the actual number of patients with the term. To account for this we adjust each probability by a bias correction factor such that the sum of the adjusted patient probabilities is equal to the actual number of patients with the term. The exact form of this correction factor is given in the supplemental “Math.pdf”.

A limitation of logistic regression is the lack of a confidence metric on each predicted probability output by the regression model. For instance, predicted probabilities might be more accurate for patients of western European than African ancestry, because the corpus data is skewed for this demographic variable (see Table S1). To overcome this limitation, we divide our data into 6 randomized partitions, balanced so that the number of affected individuals in each partition is approximately equal. This partitioning is accomplished using “StratifiedKFold” from the python package sklearn^46^. Next each partition is used to fit a logistic regression model, using the previously determined regularization strength. Each model is used to predict the probability of each person having the term. These 6 probabilities are used to calculate a sample variance for each person-term probability, 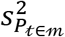, where 𝓂 represents a patient’s medical record, 𝓉 represents a medical term, and *P*_𝓉∈𝓂_ is the probability of term 𝓉 in 𝓂. To be clear these 6 LRMs are only used to calculate sample variance, while the LRM trained on the full dataset is used per-patient per-term probability predictions.

### Poisson Binomial for term pair p-values

Our null hypothesis is that pairs of medical terms are independently distributed in University of Utah medical records, *H*_0_: 𝓉1 ⊥ 𝓉2|ℳ. A significant p-value would indicate that a pair of terms co-occur more often than would occur by chance. Given two independent terms, 𝓉1 ⊥ 𝓉2 the probability the two would occur by chance in a given person’s medical record 𝓂, is the product of their individual probabilities:

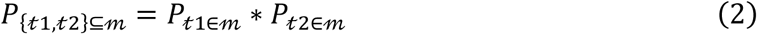

A naïve approach assumes person-term probabilities are equal to population incidence rates:

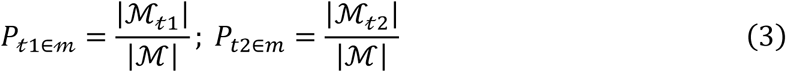

Using the naïve approach, person-term-pair probabilities follow the binomial distribution with

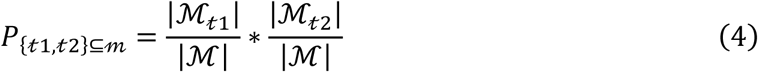

However, using logistic regression, we have different probabilities for each person/term pair. The Poisson binomial distribution is the discrete distribution of a sum of Bernoulli trials where the probability of each trial differs. Thus, using logistic regression, our data follows a Poisson binomial distribution.

Because the cumulative distribution function (CDF) of a Poisson binomial is computationally tractable only for a small number of values, numerous approximations have been developed^52^. We use the normal approximation because it is fast and accurate for large datasets. To determine a p-value using the normal approximation, the mean and variance for the Poisson binomial are calculated and used as parameters for a normal distribution. The mean of a Poisson binomial represents the expected number of University of Utah patients who will have in their medical record both terms in the pair and is calculated as the sum of probabilities for each person 𝓂:

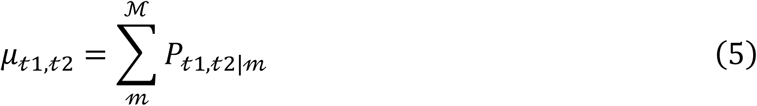

The variance of a Poisson binomial is likewise similar in form to a binomial distribution:

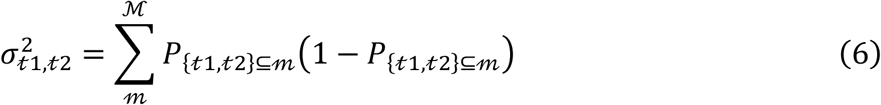

The Poisson binomial variance is augmented with the logistic regression variances described in the previous section 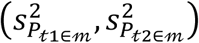 using the product rule and the law of total variance. One can think of these variances as *measurement error* for *P*_𝓉1⊆𝓂_ and *P*_𝓉2⊆𝓂_ and they are larger for rare terms.

The probability that a person has a pair of terms, *P*{_𝓉1,𝓉2_}⊆𝓂, can be so rare it exceeds the limits of floating-point arithmetic. Thus, we implement our methods in log-space. Our logistic regression models report per-patient per-term log probabilities. We calculate *ln*(*μ*_𝓉1,𝓉2_) and 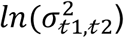 rather than summing in normal space. We use a numerical approximation to calculate the *ln*(*pvalue*) of a normal distribution^53^ (implemented as “gsl_sf_log_erfc” in the Gnu scientific library^54^). To report significance, we use throughout this paper an alpha threshold of 0.05. Since we calculate p-values for 4,622,320 pairs of medical terms, our Bonferroni corrected alpha is set to 1.08e-08.

### Direction P-values

Given two terms that occur together in medical records more often than would occur by chance, which term tends to occur first in the medical record, or do they tend to occur in the same time frame? We calculate p-values for the temporal nature of each association. For each patient with medical record *m*, the date of the first occurrence of each term in *m* is recorded. Pairs of terms occurring within a window of size W are labeled as “in-window”. Pairs of terms occurring outside of W contribute to the 𝓉1 → 𝓉2 count or the 𝓉2 → 𝓉1 count. For the analyses presented here, we chose a 90-day window, as this duration decreases noise associated with the date of information capture within the medical record, but the approach is valid over any interval. Note that some term relationships – such as a surgery procedure followed by an infection diagnosis – will only show a significant direction with a window smaller than 90 days. PBC-Web includes 4 window sizes (30, 90, 365, 730).

For a person with medical record 𝓂, containing terms 1 and 2 ({𝓉1, 𝓉2} ⊆ 𝓂), the probability that term 1 occurs before term 2 is a function of the ratio of the probabilities of the 2 terms:

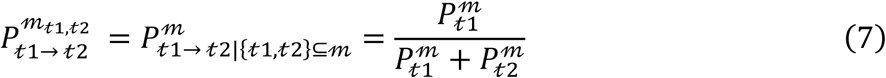

Given {𝓉1, 𝓉2} ⊆ 𝓂, the probability 𝓉1 and 𝓉2 occur within a time window of size *W*, is a function of the span or length of a person’s medical history, *Span_m_*:

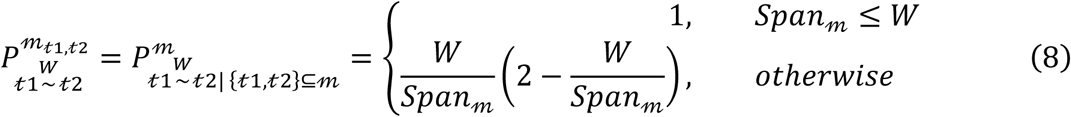

The above formula represents the percent of timepoints *t*1 and *t*2 that fall within W days of each other. The derivation of the formula is given in the supplemental “Math.pdf”.

The product of (7) and (8) gives the probability term 1 would precede term 2 within a window of size W:

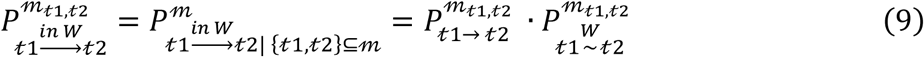

Similar logic can be applied to derive the “out-of-window” probability of term 1 occurring at least W days before term 2. A more complete explanation is found in supplemental “Math.pdf”. Direction p-values are calculated using a normal approximation of the Poisson Binomial CDF as in the previous section.

### Code, web-development, and calculations

We implement our statistical analysis using Python, Cython and C. Cython is a static compiler for Python and the extended Cython programming language^55^. Scikit-learn^51^ was used for Logistic regression studies. All the figures accompanying this article were generated using Matplotlib^56^. Our website is built using the Flask web framework^57^. The backend is pure python and the front end is JavaScript and D3^58^. Logistic regression modeling and pairwise calculation of Poisson Binomial p-values were performed at the University of Utah Center for High Performance Computing (CHPC) PHI protected environment. Training 3041 logistic regression models while tuning the regularization strength with cross validation took 1959 CPU hours. Maximum memory usage was 174 MB. Calculating comorbidity statistics for 4,623,841 term pairs took 7455 CPU hours while maximum memory usage was 87 MB.

## Supporting information

Math.pdf

## Data Availability

In this paper we calculate comorbidity statistics for all pairs of medical billing codes - including diagnoses, procedures, and medications. All of these p-values are available to query and download from the following link:
https://pbc.genetics.utah.edu/lemmon2021

## Acknowledgements

The following collaborators have provided valuable discussion, feedback, and insight which has guided development of PBC: Bruce Bray, Vikrant Deshmukh, Karen Eilbeck, Edgar Javier Hernandez, Rashmee Shah. We thank members of the University of Utah EDW for facilitating access to medical records. The computational resources used were partially funded by the NIH Shared Instrumentation Grant 1S10OD021644-01A1.

This research was supported by the AHA Children’s Strategically Focused Research Network grant (17SFRN33630041) and the Nora Eccles Treadwell Foundation. Gordon Lemmon was supported by NRSA training grant T32H757632. Sergiusz Wesolowski was supported by NRSA training grant T32DK110966-04 and the AHA Children’s Strategically Focused Research Network Fellowship award (17SFRN33630041).

## Author Contributions

Gordon Lemmon is the senior research associate leading PBC development and validation. Sergiusz Wesolowski is an applied mathematician who has helped formalize our approach to statistical testing. Alex Henrie was a software engineer on the project. Martin Tristani-Firouzi and Mark Yandell conceived of the project and secured research funding and played a key role in scientific discussions regarding development of PBC. All authors edited the manuscript.

## Data Availability

In this paper we calculate comorbidity statistics for all pairs of medical billing codes – including diagnoses, procedures, and medications. All of these p-values are available to query and download from the following link: https://pbc.genetics.utah.edu/lemmon2021.

## Code Availability

We provide a CodeOcean capsule with code and data; the link is submitted by the editor to the reviewers during the peer-review process.

## Supplemental Display Items

**Figure S1.**
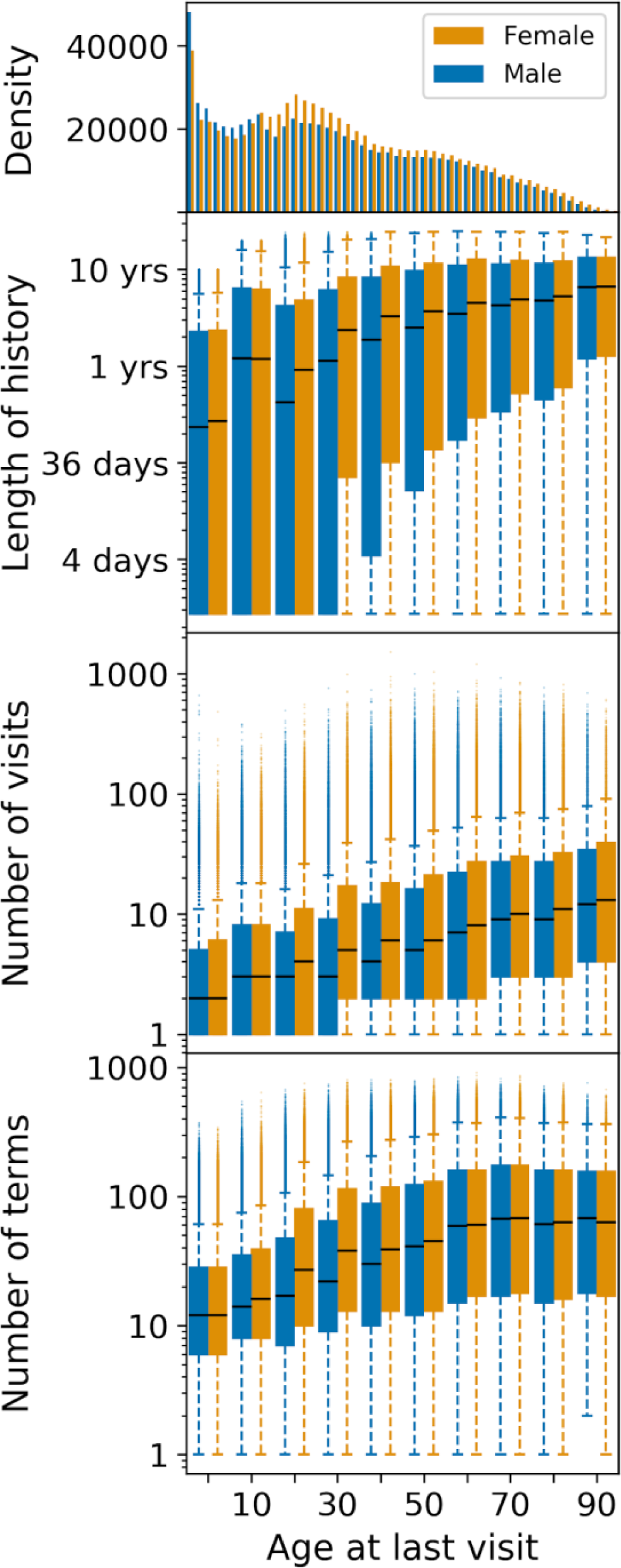
University of Utah medical records binned by age-decade. Boxplots show median (black line), 25^th^ and 75^th^ percentile (box ends), 95^th^ and 5^th^ percentile (whisker caps) and outliers. Number of terms (bottom panel) is a count of distinct diagnoses, procedures and medications found in each patient’s medical history.

**Figure S2.**
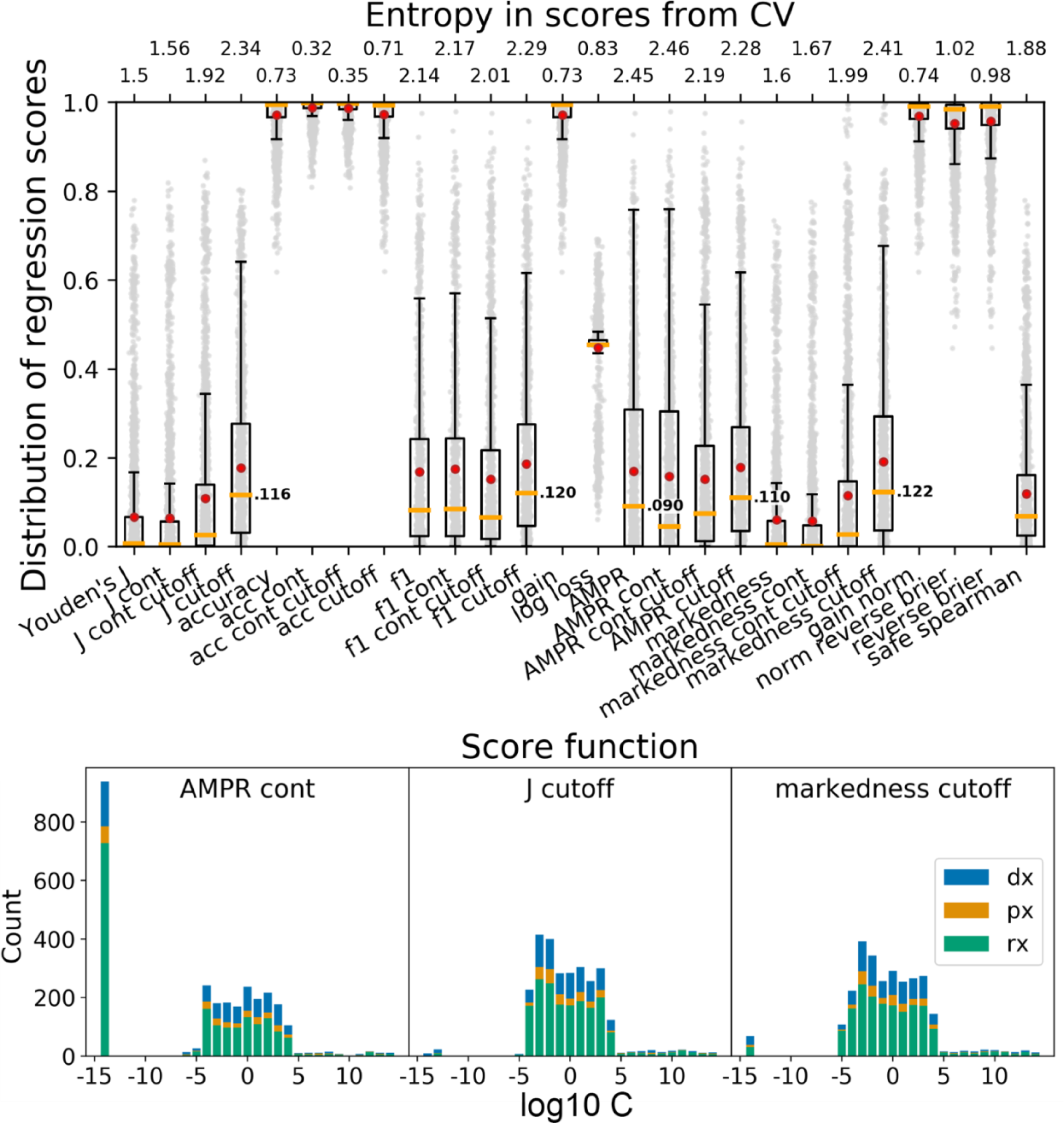
J cutoff maximizes entropy and minimizes outliers. Comparison of score functions for logistic regression C-value optimization. For each score function, we evaluated C-values ranging from 10-14 to 10^14^. Top: For each of 3041 DX, PX, and RX terms, we use cross validation to select the C-value that achieves the best score. Each boxplot contains these 3041 best scores as evaluated with different score functions. Bottom: Distribution of C-values for 3 score functions with high entropy. J_cutoff was chosen for downstream analysis because it has high entropy and has a smooth C-value distribution without the large outlier at C=-14.

**Figure S3.**
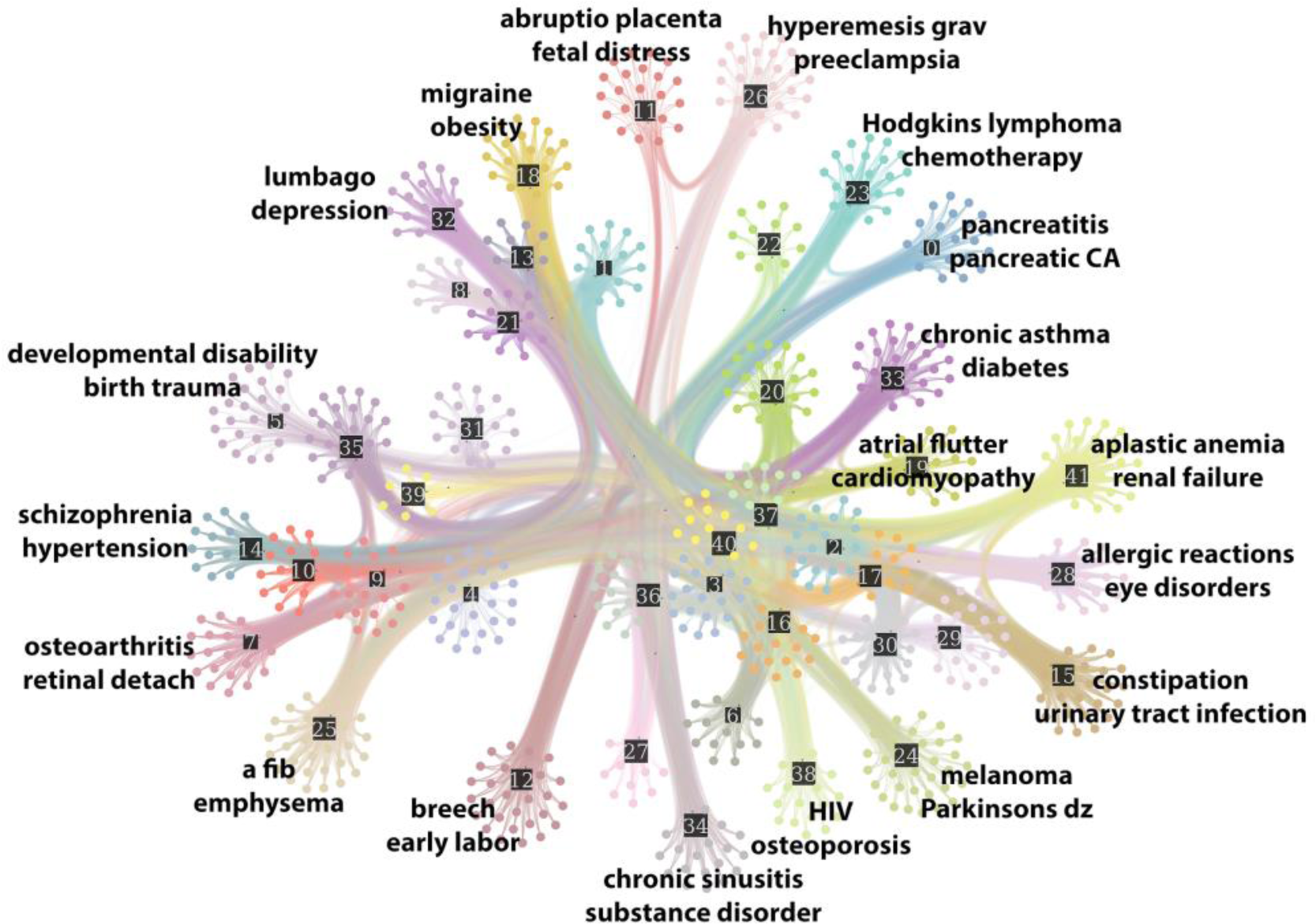
Minimum description length of the comorbidity network discovered by the PBC approach for diagnoses in the University of Utah EDW. Examples of significantly associated medical conditions within each cluster are displayed. Citations supporting these associations are listed in Suppl Table S5.

**Figure S4.**
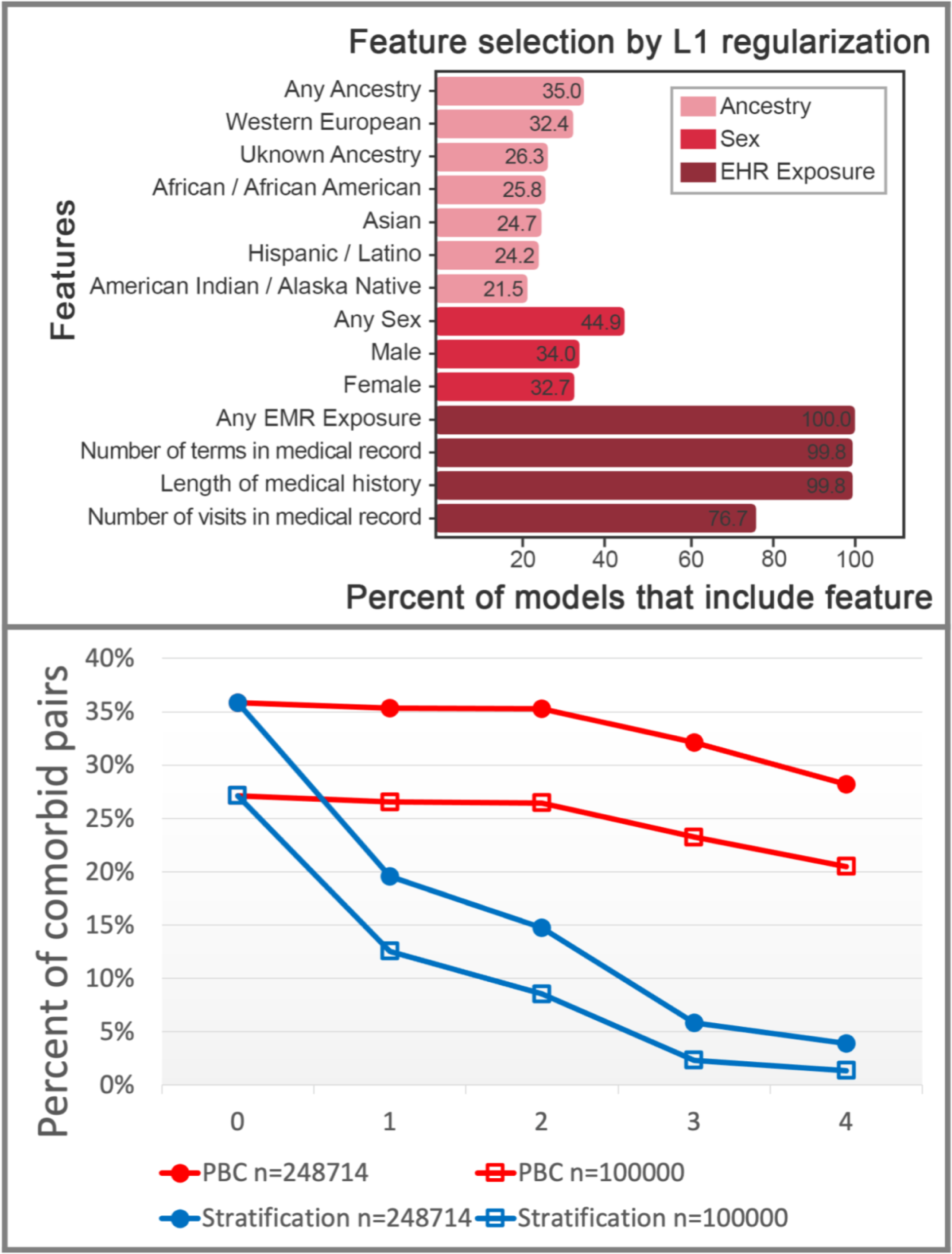
Deployment of PBC on MIMIC-IV EHR data. See Figure 1 legend for description of top panel and Figure 2 legend for description of bottom panel. Bottom panel, the X-axis ticks correspond to the addition of regression features (PBC) or stratification criteria from left to right: 0 - no features, no stratification, 1- gender/female, 2 - ancestry/African American, 3 - length of medical history/at least 2 years, 4 - number of visits/at least 3 visits. The MIMIC-IV results are very similar to the University of Utah results, reinforcing a key message of this paper - that PBC retains the power to identify comorbid relationships that are lost by stratification.

**Table S1.** University of Utah patient demographics. Total number of patients is 1,604,818.

| Gender |  | Ethnicity |  |
| --- | --- | --- | --- |
| Female | 868758 | Not hispanic | 894511 |
| Male | 815855 | Hispanic or latino | 176944 |
| Unknown | 192 | Unknown | 613350 |
| Ancestry |  |  |  |
| Caucasian | 959848 | Asian | 30952 |
| Unknown/other | 644805 | African American | 25074 |
| Native Hawaiian or Pacific Islander | 14445 | American Indian or Alaska Native | 9681 |
| Financial Class (insurance) |  |  |  |
| Commercial | 912535 | Medicaid/uninsured | 379651 |
| Medicare | 159668 | Other/Unknown | 152,964 |

**Table S2.**
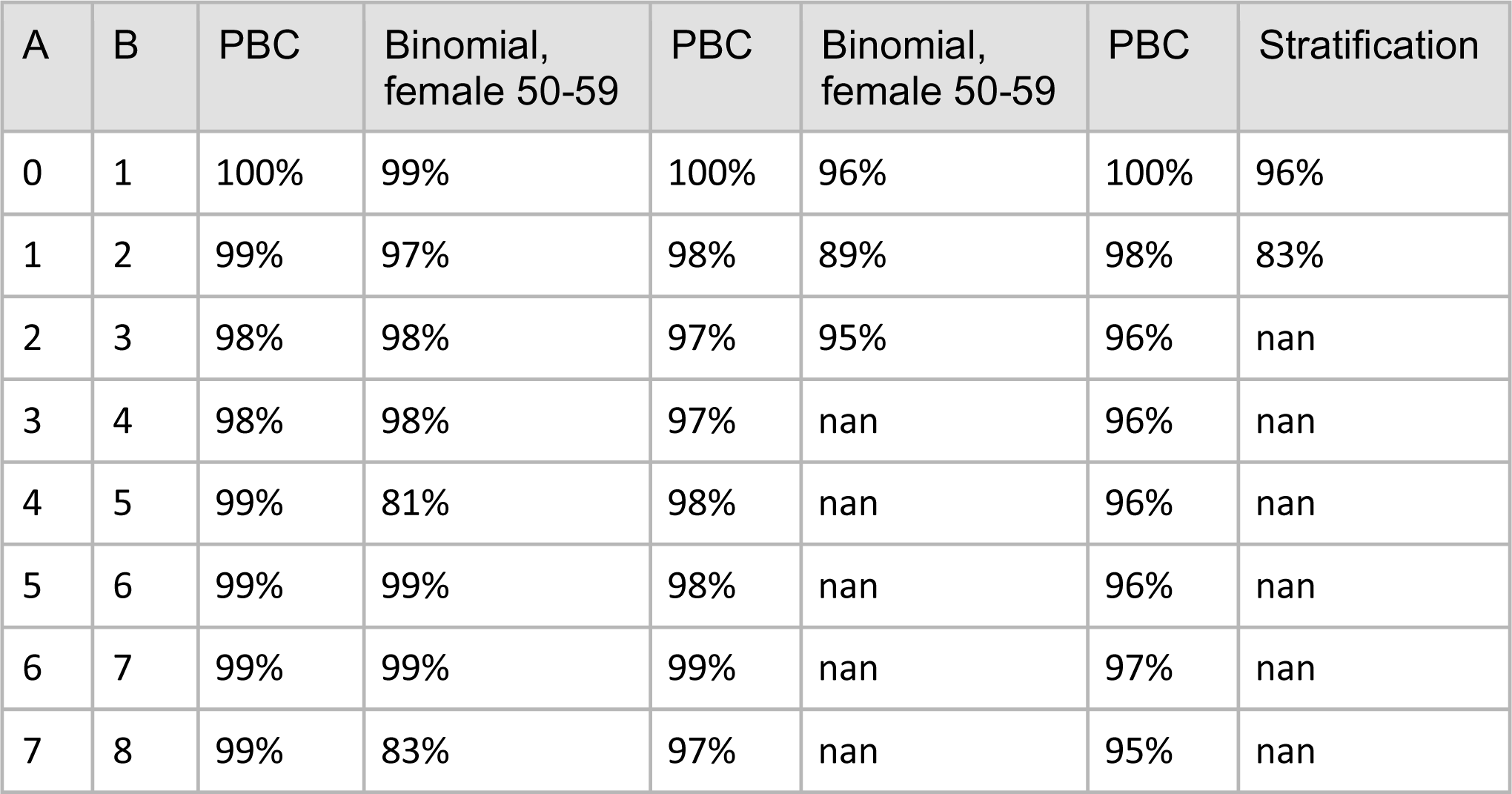
Overlap between comorbidities plotted in Figure 2. Each line in Figure 2 consists of 9 points. For every two adjacent points in these lines, we calculate the size of the intersection of the left point (A) and the right point (B) and divide by the size of B. In set notation, we calculate |A∩B| / |B|, i.e., the percent of B that is contained in A. As seen in this table, the comorbidities discovered as features are added - are almost entirely subsets of the model without the feature. Values of nan are present when B is empty.

**Table S3.** PBC retains known breast cancer comorbidities lost by the stratification approach. Comorbidities passing a Bonferroni corrected alpha threshold of **1.08e-8** are colored blue. Stratification by sex and age (considering only females in their 50s) eliminates significant associations for most known comorbidities of breast cancer, with the exception of endometriosis, a comorbidity of uncertain significance. PBC retains statistical power while modelling the effects of confounding variables. Our approach does not consider endometriosis as a comorbid condition of breast cancer.

| Representative Comorbidities of Breast Cancer: p-values |  |  |  |
| --- | --- | --- | --- |
| Potential Comorbidity | binomial all data | binomial, female 50-59 | PBC |
| Hypertension | 1e-1680 | 3.2e-8 | 1e-63 |
| Osteoarthritis | 1e-1491 | 2.5e-5 | 1e-101 |
| Cancer of colon | 1e-265 | 1.3e-6 | 1e-25 |
| Melanomas of skin | 1e-165 | 1.6e-8 | 1e-15 |
| Cancer of kidney | -1e-108 | 5e-4 | 1.6e-13 |
| Endometriosis | 1e-106 | 1e-15 | 3.2e-6 |

**Table S4.** Replication of Table 1 on MIMIC-IV data. See legend for Table 1. MIMIC-IV data does not include patient age, ethnicity or insurance type so these rows are omitted. Only 1 patient in the MIMIC-IV data set has both multiple myeloma and multiple sclerosis. For the other two known comorbidites - the trend is clear - PBC retains power as additional features are added to the model. Stratification results in a loss of statistical power.

| Stratification filters | PBC features | Concussion and migraine |  | Multiple myeloma and Multiple sclerosis |  | Cancer of pancreas and hypertension |  |
| --- | --- | --- | --- | --- | --- | --- | --- |
|  |  | P-values |  |  |  |  |  |
| | | $\chi^2$ | PBC | $\chi^2$ | PBC | $\chi^2$ | PBC |
| No filters (n=248,714) | none | -2.46 | -2.46 | -0.00 | -0.00 | -420 | -420 |
| female (n=128676) | +sex | -2.43 | -2.46 | -0.00 | -0.00 | -183 | -419 |
| +Caucasian (n=78092) | +ancestry | -1.50 | -3.02 | -0.00 | -0.00 | -117 | -391 |
| +3yr history (n=10239) | +span | 0.0 | -2.38 | -0.00 | -0.00 | -1.88 | -331 |

**Table S5.** Replication of Table 2 on MIMIC-IV data. See legend for Table 2. While the dataset is too small to measure a significant comorbidity between Sickle Cell Anemia and Malaise and Fatigue using either method, we still see that PBC retains statistical power lost by stratification. For very small sample sizes, stratification is not an option.

| Stratum filters | PBC features | Sickle Cell Anemia paired with Malaise and Fatigue |  |  |
| --- | --- | --- | --- | --- |
| | | Stratum Pair count | $\chi^2$ p-value | PBC p-value |
| No filters (n=477,070) | no features | 9 | -2.00 | -2.00 |
| Caucasian (n=276,496) | ancestry | 0 | 0 | -0.97 |
| African American (n=7,035) | ancestry | 8 | -0.67 | -0.97 |
| +Female (n=2,789) | +gender | 5 | 0 | -1.5 |

**Table S6:** Several co-occurrence and out-of-window p-values identified by PBC. Every p-value shown below passes a Bonferroni corrected alpha threshold of **1.08e-8**. Each pair of terms exhibits a significant temporal trend as indicated by arrows.

| T1 | T2 | Comorbid log10 p-value | Out-of-window Direction | Directional log10 p-value |
| --- | --- | --- | --- | --- |
| Milrinone (1,676) | Heart Transplant (122) | 1e-104 | → | 1e-32 |
| T2D (97,484) | CKD (15,123) | 1e-3374 | → | 1e-195 |
| HIV (2,730) | Pneumocystis (203) | 1e-120 | → | 1e-16 |
| Obesity due to excess calories (26494) | Essential Hypertension (-1299) | 1e-848 | → | 1e-1299 |

**Table S7:** Citations corresponding to comorbidities discovered by minimum description length clustering. See Figure S3 for a graphical representation of MDL clusters.

| Term 1 | Term 2 | PubMed ID |
| --- | --- | --- |
| abruptio placenta | fetal distress | 26393335 |
| hyperemesis gravidarum | preeclampsia | 23360164 |
| Hodgkins lymphoma | chemotherapy | 26541251 |
| pancreatitis | pancreatic cancer | 30315287 |
| Chronic asthma | diabetes | 30489598 |
| aplastic anemia | renal failure | 17426071 |
| allergic reactions | eye disorders | 31343437 |
| constipation | urinary tract infection | 30212423 |
| melanoma | Parkinson's disease | 29991141 |
| HIV | osteoporosis | 25709813 |
| chronic sinusitis | substance disorder | 28812909 |
| breech | early labor | 31741046 |
| atrial fibrillation | emphysema | 25900353 |
| osteoarthritis | retinal detachment | 20462780 |
| schizophrenia | hypertension | 27855222 |
| developmental disability | birth trauma | 31240076 |
| lumbago | depression | 31703727 |
| migraine | obesity | 27358118 |

