## Supplementary material for "A Poisson binomial based statistical testing framework for comprehensive comorbidity discovery across massive Electronic Health Record datasets": Math.pdf

### DERIVATION OF CO-OCCURRENCE AND DIRECTION P-VALUES

#### BACKGROUND

Let $\mathcal{M}$ represent a set of medical records

$$|\mathcal{M}| = \ cardinality\ of\ \mathcal{M}$$

Let $m \in \mathcal{M}$, represent a medical record chosen from $\mathcal{M}$

A medical record $m$ consists of one patient and all of their interactions or encounters with University of Utah hospitals and clinics. Each encounter consists of a date and a list of medical terms.

A medical term $t$ can represent any diagnosis, procedure, or medication. Let $\mathcal{T}_m$ represent the set of terms in all encounters in $m$.

$$\mathcal{T}_m = \{t \in m\}$$

Let $\mathcal{T}$ represent the set of all terms in all encounters in $\mathcal{M}$ and $t \in \mathcal{T}$, represent a term chosen from $\mathcal{T}$,

$$\mathcal{T} = \left\{t \middle| \underset{m \in \mathcal{M}}{\exists} : t \in \mathcal{T}_m\right\}$$

Let $\mathcal{M}_t$ denote the set of all medical records containing $t$

$$\mathcal{M}_t = \left\{m \middle| \underset{m \in \mathcal{M}}{\exists} : t \in m\right\}$$

$$m_t = an\ \ m\ \ chosen\ from\ \mathcal{M}_t$$

Let $\mathcal{M}_{t1,t2}$ represent the set of medical records containing terms $t1$ and $t2$

$$\mathcal{M}_{t1,t2} = \left\{m \middle| \underset{m \in \mathcal{M}}{\exists} : \{t1, t2\} \subseteq m\right\}$$

$$m_{t1,t2} = an\ \ m\ \ chosen\ from\ \mathcal{M}_{t1,t2}$$

Let $t1 \rightarrow t2$ indicate that $t1$ occurs before $t2$.

$$\mathcal{M}_{t1\rightarrow t2} = \left\{m \middle| \underset{m \in \mathcal{M}_{t1,t2}}{\exists} : t1 \rightarrow t2\right\}$$, i.e. set of records where $t1$ occurs before $t2$

Let $W$ represent a window (i.e. a time interval) measured in days, within which two terms are considered “co-occurring”. If two terms are “co-occurring” within the time interval $W$ we write: $t1 \overset{W}{\sim} t2$. If two terms are present in a medical record but not co-occurring in $W$ then we write: $t1 \overset{W}{\nsim} t2$. Similarly, $t1 \overset{W}{\rightarrow} t2$ indicates that term 1 occurs at least $W$ days before term 2. Thus, we can define:

$$\mathcal{M}_{t1 \overset{W}{\sim} t2} = \left\{m \middle| \underset{m \in \mathcal{M}_{t1,t2}}{\exists} : t1 \overset{W}{\sim} t2\right\},$$

i.e. set of records where $t1$ and $t2$ occur within $W$, and

$$\mathcal{M}_{t1 \overset{W}{\rightarrow} t2} = \left\{m \middle| \underset{m \in \mathcal{M}_{t1\rightarrow t2}}{\exists} : t1 \overset{W}{\rightarrow} t2\right\},$$

i.e. set of records where $t1$ occurs more than $W$ days before $t2$

#### LOGISTIC REGRESSION TO DETERMINE PER PERSON TERM PROBABILITIES

##### NOTATION

For brevity we introduce the following notation:

$$P_t^m = P_{t \in m}$$

$$P_{t1,t2}^m = P_{\{t1,t2\} \subseteq m}$$

##### DESCRIPTION OF LOGISTIC REGRESSION MODELING

For each $t$ in $\mathcal{M}$, we use logistic regression to determine a best fit equation for the probability of seeing $t$ in $m$ based on the following parameters: (1) age of $m$ at last visit; (2) length of medical history for $m$; (3) number of visits, $|\mathcal{V}_m|$; (4) number of terms in $m$; (5) gender of $m$; (6) race of $m$; (7) ethnicity of $m$; (8) financial class of $m$. The outcome is an equation for each term $t$, which predicts the probability of $t$ in $m$. In the equation below, $x_1$ through $x_8$ represent the features of $m$ as listed above.

$$P_t^m = \frac{1}{1 + exp\big(-(\beta_0 + \beta_1 x_1 + \cdots + \beta_8 x_8)\big)}$$

L2 penalized logistic regression approach minimizes the following cost function using a coordinate descent [1]:

$$\min_{\beta,c} \frac{1}{2} \|\beta\|_2 + C \sum_{i=1}^{n} log\left(exp\left(-y_i(X_i^T \beta + c)\right) + 1\right)$$

The value of $C$ represents the inverse of regularization strength. Smaller values represent stronger regularization. Regularization is the application of a penalty when increasing the effect of input features on output probability, in order to reduce overfitting.

We sought to determine the optimal C-value empirically. We did so using stratified 3-fold cross-validation. By stratified, we mean that the average number of positive cases $|\mathcal{M}_t|$ in each fold is approximately equal. We cross validate using each of the C-values in the set $\{10^{-14}, 10^{-13}, 10^{-12}, \ldots, 10^{12}, 10^{13}, 10^{14}\}$ to determine the C-value that optimizes our chosen score function.

#### CHOOSING A CROSS VALIDATION SCORE FUNCTION AND OPTIMIZING “C”

The score function generally used by cross-validation to evaluate the performance of the trained model on the left-out test data is mean accuracy. However, for our dataset, mean accuracy entirely fails to discriminate among competing logistic regression models. To explain why, consider rarity of medical terms. For instance, in our data $|\mathcal{M}| \cong 1.7\ million$ and $|\mathcal{M}_{tuberculosis}| = 206$. The probability of any given person having tuberculosis is small, but might be several orders of magnitude different than another person’s probability based on age, gender, race, etc. Since even the most at-risk population has probabilities close to zero, logistic regression models always classify individuals as unaffected.

We considered a variety of standard and custom score functions as described in the next section. Instead of scoring based on correct or incorrect classification (0 or 1) – “continuous” score functions score as the difference between the correct classification and the predicted probability from logistic regression. Instead of using 50% predicted probability as the threshold for classification, “cutoff” score functions use the threshold value that leads to a number of positive classifications equal to that found in the training data.

Below is a graph showing the distribution of 3-fold cross-validation scores using various scoring functions. Each column represents 2784 logistic regression cross validation scores, carried out for each of 2784 CCS procedure or CCS diagnosis codes and RXNorm medication codes. Each logistic regression includes 1.6 million medical records from the University of Utah.

Yellow lines represent the median, the boxes extend from the lower quartile to the upper quartile of the data, whiskers extend to 1.5 times the interquartile range beyond the box. Red dot represents the mean of the data. Outliers are black with a transparency of 0.5.

With the exception of log loss, a value of 1 indicates a perfectly accurate prediction. For log loss, zero indicates a perfect prediction. Score functions with a mean near one or zero each suffer from the same problem explained above in relation to mean accuracy - the true positives signal is lost amidst the abundance of negative outcomes. This also applies to log loss.

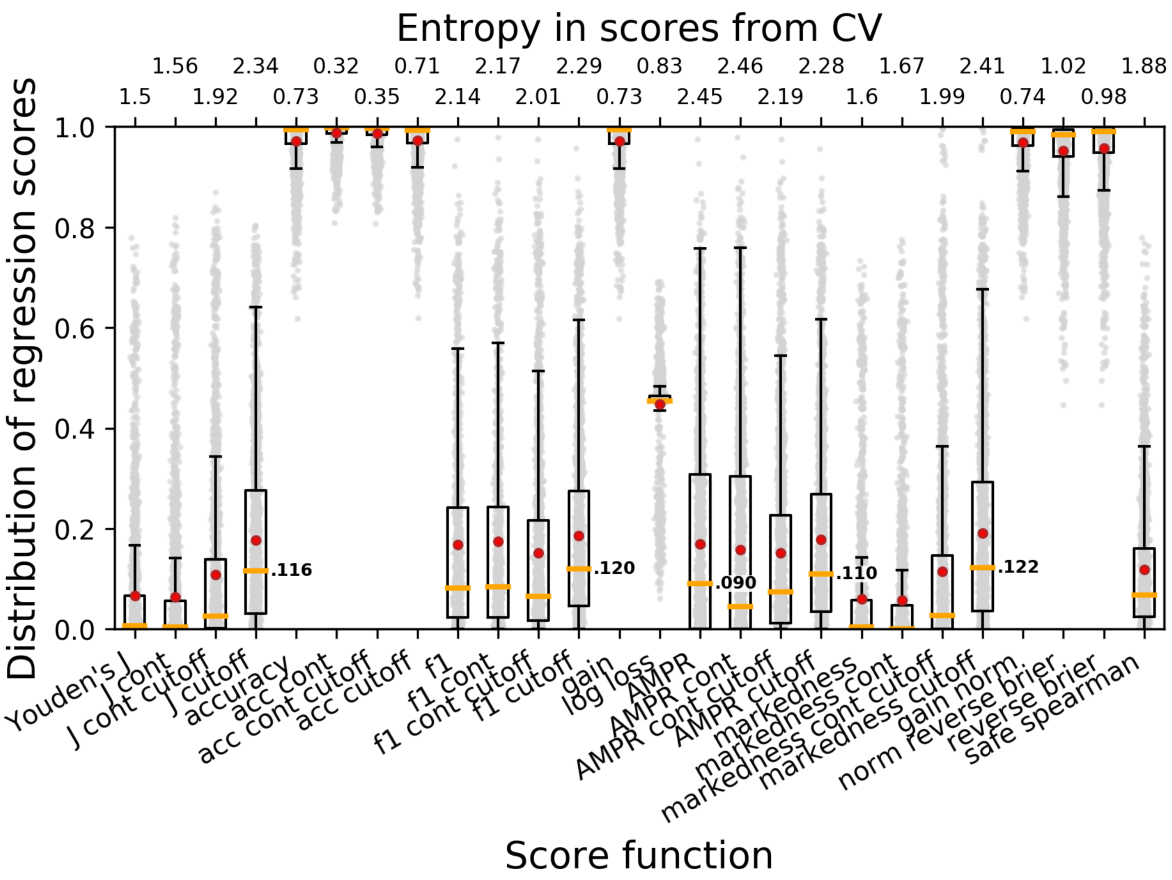

#### DESCRIPTION OF SCORE FUNCTIONS EVALUATED FOR USE WITH CROSS VALIDATION

$$f_{t,m} = forcast\ or\ predicted\ probability\ of\ term\ t\ in\ medical\ record\ m$$

$$o_{t,m} = observation\ of\ whether\ term\ t \in m, takes\ a\ value\ of\ 0\ or\ 1$$

$$TP = number\ of\ true\ positives$$

$$TN = number\ of\ true\ negatives$$

$$FP = number\ of\ false\ positives$$

$$FN = number\ of\ false\ negatives$$

$$TPR = \frac{TP}{TP + FN}\ (aka.\ true\ positive\ rate; sensitivity; or\ recall)$$

$$TNR = \frac{TN}{TN + FP}\ (aka.\ true\ negative\ rate; specificity; or\ selectivity)$$

$$PPV = \frac{TP}{TP + FP}\ (aka.\ positive\ predictive\ value; or\ precision)$$

$$J = informedness = sensitivity + specificity - 1 = \mathrm{TPR} + \mathrm{TNR} - 1$$
$$(aka.\ \text{Youden's J statistic})$$

$$acc = \frac{TP + TN}{TP + TN + FP + FN}\ (aka.\ accuracy)$$

$$markedness = PPV + NPV - 1$$

$$f1 = HMPR = 2 \cdot \frac{precision \cdot recall}{precision + recall} = 2 \cdot \frac{PPV \cdot TPR}{PPV + TPR}$$

$$(aka. harmonic\ mean\ of\ precision\ and\ recall)$$

$$AMPR = \frac{precision + recall}{2} = \frac{PPV + TPR}{2}$$

$$(aka. arithmetic\ mean\ of\ precision\ and\ recall)$$

$$BS_t = brier\ score = \frac{1}{|\mathcal{M}|} \sum_{m}^{\mathcal{M}} \left(f_{t,m} - o_{t,m}\right)^2 ,$$

$$gain_t = \frac{1}{|\mathcal{M}|} \sum_{m}^{\mathcal{M}} f_{t,m} - o_{t,m}$$

$$LL_t = log\_loss = -\frac{1}{|\mathcal{M}|} \sum_{m}^{\mathcal{M}} o_{t,m} \cdot log\, f_{t,m} + \left(1 - o_{t,m}\right) \cdot log\left(1 - f_{t,m}\right) ,$$

$$\rho = Spearman's\ rank\ correlation$$

$$achieves\ a\ value\ of\ 1.0\ when\ all\ o_{t,m} = 0\ precede\ o_{t,m} = 1\ after\ sorting\ by\ f_{t,m}$$

#### SELECTING A SCORE FUNCTION

A score function that spreads out the scores for different logistic regression models has the most power to distinguish informative from uninformative models. Thus, entropy was chosen as the metric for selecting a score function for later use with cross validation. As shown in figure above, the 3 functions with the highest entropy were J cutoff, AMPR, and AMPR cont. Below is a plot of the distribution of C-values for each per-term logistic regression, for each score function:

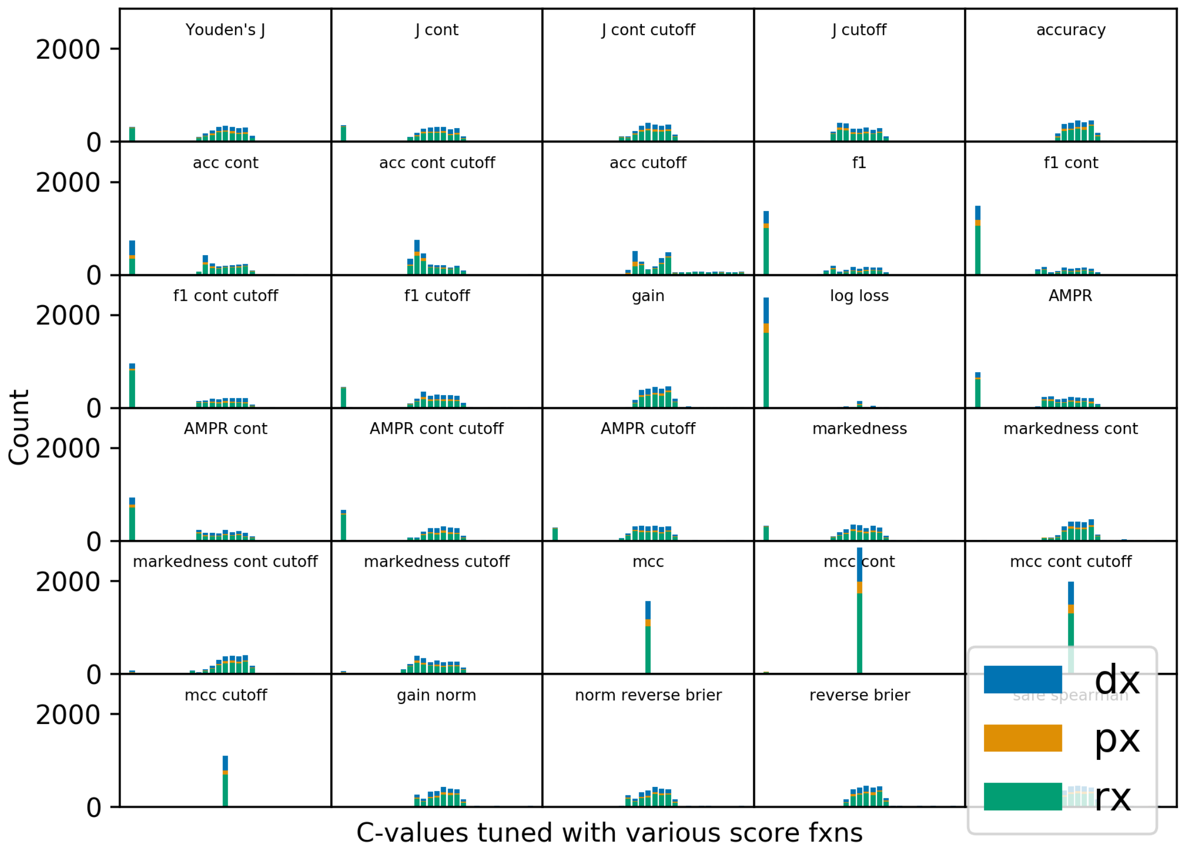

Many of the score functions evaluated suffer from a large outlier at the lowest tested C-value of 1e-14. This C-value effectively turns off regression entirely, setting coefficients to zero. Of the aforementioned high entropy score functions under consideration, AMPR and AMPR cont suffer from a large outlier at 1e-14. Therefore, we chose J cutoff as our preferred score function for optimizing C.

#### VARIANCE FOR LOGISTIC REGRESSION PROBABILITY PREDICTIONS

A limitation of standard logistic regression is that the probabilities produced by the decision function do not include a confidence interval. We overcame this limitation by training multiple logistic regression models on non-overlapping subsets of the data and calculating sample variance from the results of multiple decision functions. The result is a separate variance on the prediction of each term occurring in each person.

For each term we stratify the data into 6 subsets with equal representation of the term. We train 6 logistic regression models using the predetermined optimal C value. We calculate unbiased sample variance using the 6 values of $P_t^m$ produced by 6 logistic regression models:

$$s^2_{P_t^m} = UnbiasedSampleVariance(P_t^m) = \frac{1}{n-1}\sum_{i=1}^{6}(P_t^m)_i - \overline{P_t^m}^2$$

#### NORMALIZATION OF LOGISTIC REGRESSION PROBABILITIES

Logistic regression models are used to calculate the probability that a given patient has a given term in his or her medical record. It is often the case that the probabilities output by the trained model do not sum up to the total number of patients with the given term in their medical record. When logistic regression models exhibit this bias, we apply a bias correction factor to each patient probability:

$$\grave{P}_t^m = P_t^m * \frac{|\mathcal{M}_t|}{\sum_m^{\mathcal{M}} P_t^m}$$

such that the new probabilities sum to equal the actual term count:

$$\sum_m^{\mathcal{M}} \grave{P}_t^m \equiv |\mathcal{M}_t|$$

For clarity below, we omit the accent mark, but in practice we always use $\grave{P}_t^m$ rather than $P_t^m$.

### CO-OCCURANCE P-VALUES

The goal of this analysis is to determine which medical terms occur together more often than expected. To this end, we calculate a co-occurrence p-value for every pair of medical diagnoses, procedures, and medications. This involves calculating expected value and variance from a Poisson-binomial distribution.

#### CALCULATION OF PAIRWISE EXPECTATION AND VARIANCE

The probability of two independent terms co-occurring in $m$ is simply:

$$P_{t1,t2}^m = P_{t1}^m \cdot P_{t2}^m$$

Our goal is to test whether this assumption of independence holds true. Since the observation of $t_1$ and $t_2$ co-occurring in $m$ is a Bernoulli trial, the variance on $P_{t1,t2}^m$is expressed as:

$$\sigma^2_{P_{t1,t2}^m} = Var\left(P_{t1,t2}^m\right) = P_{t1,t2}^m\left(1 - P_{t1,t2}^m\right)$$

and each $m$ has a different $P_{t1,t2}^m$, the number of records in $\mathcal{M}$ that have terms $t_1$ and $t_2$ is modeled using a Poisson-Binomial distribution. If $\pi$ represents the vector of probabilities $P_{t1,t2}^m$ for all $m$, then:

$$\left|\mathcal{M}_{t1,t2}\right| \sim PoiBin(\pi)$$

The expected value for the number of records in $\mathcal{M}$ that have terms $t1$ and $t2$:

$$\mu_{|\mathcal{M}_{t1,t2}|} = E\left(\left|\mathcal{M}_{t1,t2}\right|\right) = \sum_{m \in \mathcal{M}} P_{t1,t2}^{m}$$

The variance for the expected number of records in $\mathcal{M}$ that have terms $t1$ and $t2$:

$$\sigma_{|\mathcal{M}_{t1,t2}|}^{2} = Var\left(\left|\mathcal{M}_{t1,t2}\right|\right) = \sum_{m \in \mathcal{M}} \sigma_{P_{t1,t2}^{m}}^{2}$$

#### INCORPORATION OF VARIANCE FROM LOGISTIC REGRESSION

The variance of a Poisson binomial distribution is augmented with the logistic regression variances described in previously. One can think of these variances as *measurement error* for $P_{t1}^{m}$ and $P_{t2}^{m}$ and they are larger for rare terms. The product rule allows us to calculate the Logistic Regression sample variance for $P_{t1,t2}^{m}$:

$$s_{P_{t1,t2}^{m}}^{2} = (P_{t1}^{m})^{2} s_{P_{t2}^{m}}^{2} + (P_{t2}^{m})^{2} s_{P_{t1}^{m}}^{2} + s_{P_{t1}^{m}}^{2} s_{P_{t2}^{m}}^{2}$$

The law of total variance allows us to calculate the Poisson binomial variance augmented by the Logistic regression sample variance:

$$\sigma_{|\mathcal{M}_{t1,t2}|}^{2} = Var\left(\left|\mathcal{M}_{t1,t2}\right|\right) = \sum_{m \in \mathcal{M}} \sigma_{P_{t1,t2}^{m}}^{2} + s_{P_{t1,t2}^{m}}^{2}$$

#### DIRECTION P-VALUES

We calculate direction p-values in two ways – the first way considers only the subset of medical records containing terms $t_1$ and $t_2$, $\mathcal{M}_{t1,t2}$. We call these conditional direction p-values. Only if the co-occurrence p-value is significant are these p-values meaningful. We also calculate direction p-values using the complete set of medical records.

#### CALCULATION OF CONDITIONAL EXPECTATION AND VARIANCE

Given that a person's medical record $m$ has terms $t_1$ and $t_2$, the likelihood that $t_1$ occurs before $t_2$ in $m_{t1,t2}$ (assuming the direction is random) is a function of each term's separate probability.

$$P_{t1 \rightarrow t2}^{m_{t1,t2}} = P_{t1 \rightarrow t2 | \{t1,t2\} \subseteq m}^{m} = \frac{P_{t1}^{m}}{P_{t1}^{m} + P_{t2}^{m}}$$

* Note on notation: We remind the reader that $P^{m}_{t1,t2}$ represents the probability that $t1$ and $t2$ are in medical record $m$, whereas $P^{m_{t1,t2}}_{t1\rightarrow t2}$ is conditioned on $t1$ and $t2$ being present in medical record $m$, i.e. $P^{m}_{t1,t2} = 1$.

Given a direction window size $W$, a patient $m$, and the length of $m$'s medical history as $Span_m$, the probability terms $t_1$ and $t_2$occur within $W$, given $t_1$ and $t_2$ occur in $m$ is:

$$P^{m_{t1,t2}}_{\underset{t1\sim t2}{W}} = P^{m}_{\underset{t1\sim t2|\,\{t1,t2\}\subseteq m}{W}} = \begin{cases} 1, & Span_m \le W \\ \dfrac{W}{Span_m}\left(2 - \dfrac{W}{Span_m}\right), & otherwise \end{cases}$$

The proof for the above comes from calculating the fraction of the square below that is shaded:

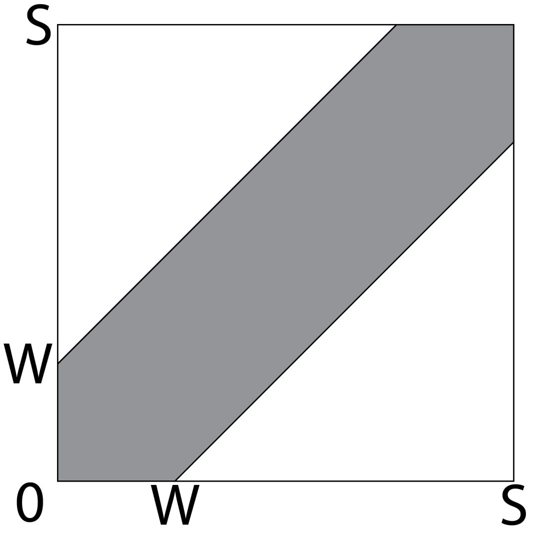

In the above square, the area of the square is $S^2$ and the area of either triangle is $\frac{1}{2}(S-W)^2$. The percent of the square that is shaded is $\frac{1-2*\frac{1}{2}(S-W)^2}{S^2}$ and algebra reduces this to the above stated equation.

The probability that $t1$ and $t2$ occur outside of $W$ is:

$$P^{m_{t1,t2}}_{\underset{t1\nsim t2}{W}} = 1 - P^{m_{t1,t2}}_{\underset{t1\sim t2}{W}}$$

The probability that term $t1$ occurs within $W$ days before $t2$ is a function of the frequency of the terms in the EDW.

$$P^{m_{t1,t2}}_{t1\xrightarrow{in\,W}t2} = P^{m}_{t1\xrightarrow{in\,W}t2|\,\{t1,t2\}\subseteq m} = P^{m_{t1,t2}}_{t1\rightarrow t2} \cdot P^{m_{t1,t2}}_{\underset{t1\sim t2}{W}}$$

The probability that term $t1$ occurs at least $W$ days before $t2$ is a function of the frequency of the terms in the EDW.

$$P^{m_{t1,t2}}_{t1 \xrightarrow{out\,W} t2} = P^{m}_{t1 \xrightarrow{out\,W} t2 |\, \{t1,t2\} \subseteq m} = P^{m_{t1,t2}}_{t1 \rightarrow t2} \cdot P^{m_{t1,t2}}_{t1 \overset{W}{\nsim} t2}$$

Since the above probabilities differ for each $m$, a Poisson-Binomial distribution can be used to calculate p-values. Let $\left|\mathcal{M}_{t1 \overset{W}{\sim} t2}\right|$ represent the number of medical records with terms $t_1$ and $t_2$ occuring within $W$. The expectation for number of times $t_1$ and $t_2$ occur in $W$ observed over $\mathcal{M}_{t1,t2}$ is the sum of the probabilities for each $m$:

$$E\left(\left|\mathcal{M}_{t1 \overset{W}{\sim} t2}\right|\right) = \sum_{m_{t1,t2} \in \mathcal{M}_{t1,t2}} P^{m_{t1,t2}}_{t1 \overset{W}{\sim} t2}$$

The variance for the above expectation is:

$$Var\left(\left|\mathcal{M}_{t1 \overset{W}{\sim} t2}\right|\right) = \sum_{m_{t1,t2} \in \mathcal{M}_{t1,t2}} P^{m_{t1,t2}}_{t1 \overset{W}{\sim} t2} \cdot P^{m_{t1,t2}}_{t1 \overset{W}{\nsim} t2}$$

The expected number of times $t1$ occurs within $W$ days before $t2$ in $\mathcal{M}_{t1,t2}$, and the variance for this expectation are:

$$E\left(\left|\mathcal{M}_{t1 \xrightarrow{in\,W} t2}\right|\right) = \sum_{m_{t1,t2} \in \mathcal{M}_{t1,t2}} P^{m_{t1,t2}}_{t1 \xrightarrow{in\,W} t2}$$

$$Var\left(\left|\mathcal{M}_{t1 \xrightarrow{in\,W} t2}\right|\right) = \sum_{m_{t1,t2} \in \mathcal{M}_{t1,t2}} P^{m_{t1,t2}}_{t1 \xrightarrow{in\,W} t2}\left(1 - P^{m_{t1,t2}}_{t1 \xrightarrow{in\,W} t2}\right)$$

The expected number of times $t1$ occurs more than $W$ days before $t2$ in $\mathcal{M}_{t1,t2}$, and the variance for this expectation are:

$$E\left(\left|\mathcal{M}_{t1 \xrightarrow{out\,W} t2}\right|\right) = \sum_{m_{t1,t2} \in \mathcal{M}_{t1,t2}} P^{m_{t1,t2}}_{t1 \xrightarrow{out\,W} t2}$$

$$Var\left(\left|\mathcal{M}_{t1 \xrightarrow{out\,W} t2}\right|\right) = \sum_{m_{t1,t2} \in \mathcal{M}_{t1,t2}} P^{m_{t1,t2}}_{t1 \xrightarrow{out\,W} t2}\left(1 - P^{m_{t1,t2}}_{t1 \xrightarrow{out\,W} t2}\right)$$

Conditional direction p-values are calculated using the normal approximation for the CDF of a Poisson binomial distribution as described above.

#### CALCULATION OF DIRECTION EXPECTATION AND VARIANCE

The previous section derived conditional probabilities for two terms occurring together or one before the other in a person's medical record, given the person has the two terms in their medical record. We can combine these conditional probabilities with the probability of the person having both terms in their medical record to obtain the overall probabilities of a person having a pair of terms in window or one before the other:

$$P^{m}{}_{\underset{t1 \sim t2}{W}} = P^{m_{t1,t2}}{}_{\underset{t1 \sim t2}{W}} \cdot P^{m}_{t1,t2}$$

$$P^{m}{}_{\underset{t1 \nsim t2}{W}} = P^{m_{t1,t2}}{}_{\underset{t1 \nsim t2}{W}} \cdot P^{m}_{t1,t2}$$

$$P^{m}{}_{t1 \xrightarrow{in\,W} t2} = P^{m_{t1,t2}}{}_{t1 \xrightarrow{in\,W} t2} \cdot P^{m}_{t1,t2}$$

$$P^{m}{}_{t1 \xrightarrow{out\,W} t2} = P^{m_{t1,t2}}{}_{t1 \xrightarrow{out\,W} t2} \cdot P^{m}_{t1,t2}$$
